# Spatial proteomic characterization of HER2-positive breast tumors through neoadjuvant therapy predicts response

**DOI:** 10.1101/2020.09.23.20199091

**Authors:** Katherine L. McNamara, Jennifer L. Caswell-Jin, Rohan Joshi, Zhicheng Ma, Eran Kotler, Gregory R. Bean, Michelle Kriner, Zoey Zhou, Margaret Hoang, Joseph Beechem, Jason Zoeller, Michael F. Press, Dennis J. Slamon, Sara A. Hurvitz, Christina Curtis

## Abstract

Addition of HER2-targeted agents to neoadjuvant chemotherapy has dramatically improved pathological complete response (pCR) rates in early-stage HER2-positive breast cancer. Still, up to 50% of patients have residual disease following treatment, while others are likely overtreated. Here, we performed multiplex spatial proteomic characterization of 122 samples from 57 HER2-positive breast tumors from the neoadjuvant TRIO-US B07 clinical trial sampled pre-treatment, after 14-21 days of HER2-targeted therapy, and at surgery. We demonstrate that proteomic changes following a single cycle of HER2-targeted therapy aids the identification of tumors that ultimately undergo pCR, outperforming pre-treatment measures or transcriptomic changes. We further developed and validated a classifier that robustly predicts pCR using a single marker, CD45, measured on-treatment, and show that CD45-positive cell counts measured via conventional immunohistochemistry perform comparably. These results demonstrate novel biomarkers to enable the stratification of sensitive tumors early during neoadjuvant HER2-targeted therapy with implications for tailoring subsequent therapy.

---

Human epidermal growth factor receptor 2 (HER2)-positive breast cancer accounts for 15–30% of invasive breast cancers and is associated with an aggressive phenotype ^1^. The neoadjuvant setting allows for early assessment of treatment response, and pathologic complete response (pCR) is a strong surrogate for long-term survival in HER2-positive disease ^2–4^. While the addition of HER2-targeted agents to neoadjuvant chemotherapy has dramatically improved pCR rates in early stage HER2-positive breast cancer, 40-50% of patients have residual disease after treatment ^5, 6^. Conversely, HER2 inhibition with two targeted agents and without chemotherapy can result in pCR, suggesting that it may be possible to eliminate chemotherapy in a subset of patients ^7–10^. Given the heterogeneity in response to HER2-targeted therapy ^5, 11^, identification of biomarkers of response beyond HER2 and estrogen receptor (ER) status is needed.Bulk gene expression profiling of pre-treatment samples has identified tumor characteristics (HER2-enriched intrinsic subtype, HER2 expression levels, and ESR1 expression levels ^10, 12–15^), and microenvironmental characteristics (increased immune infiltration ^13, 15–18^) that associate with pCR to HER2-targeted therapy. While pCR and long-term survival correlate strongly on an individual level ^2–4^, tumor characteristics may correlate in opposite directions with pCR and recurrence-free survival (RFS) (for example with the HER2-enriched intrinsic subtype correlating with higher pCR rate and shorter RFS), while immune characteristics may correlate in the same direction (with increased immune infiltration correlating with higher pCR rate and longer RFS) ^19^.

Because tumor cells are profiled simultaneously with both co-localized and distant stroma and immune cells, bulk expression profiling is an imperfect tool for analyzing tumor and microenvironmental changes across treatment. In particular, it is difficult to assign observed changes to specific geographic or phenotypic cell populations within the complex tumor ecosystem, where malignant tumor cells interact with fibroblasts, endothelial cells, and immune cells. Moreover, immune cells can be further divided into those that infiltrate the tumor core and those that are excluded ^20^. As of yet, how the tumor and immune microenvironment change during therapy remains poorly understood, necessitating multiplexed *in situ* profiling of longitudinal tissue samples. It is unknown whether such on-treatment *in situ* profiling might outperform pre-treatment bulk expression biomarkers in predicting HER2-targeted therapy response.

We used the GeoMx™ Digital Spatial Profiling (DSP, NanoString) technology to assay archival tissue from an initial discovery set of 28 patients with HER2-positive breast cancer enrolled on the neoadjuvant TRIO-US B07 clinical trial ^21, 22^, whose tumors were sampled pre-treatment, after 14-21 days of HER2-targeted therapy, consisting of lapatinib, trastuzumab, or both (on-treatment), and at the time of surgery after completion of combination chemotherapy with HER2-targeted therapy (post-treatment). We subsequently validated our results in an independent set of 29 patients from the same trial. DSP enables geographic and phenotypic selection of tissue regions for multiplex proteomic characterization of cancer signaling pathways and the tumor-colocalized immune microenvironment ^23, 24^. In particular, we characterized spatial heterogeneity in untreated breast tumors as well as changes in cancer signaling pathways and microenvironmental composition in matched on-treatment biopsies and post-treatment surgical samples by profiling 40 tumor and immune proteins across multiple pancytokeratin (panCK)-enriched regions per sample. On-treatment protein expression changed dramatically in tumors that went on to achieve a pCR and a classifier based on these data outperformed models based on transcriptomic data, as well as established predictors such as PAM50 subtype ^10, 14, 25^ and stromal lymphocytes (CelTIL) ^17^. This new spatial proteomic model robustly predicted treatment response in the validation cohort. Moreover, on-treatment CD45 protein expression alone predicted outcome with high accuracy – a finding that was reproduced with CD45 immunohistochemistry (IHC). Collectively, these results suggest new avenues to personalize therapy in early-stage HER2 -positive breast cancer.

## Results

### Spatial proteomic analysis of untreated HER2-positive breast tumors

Participants in the TRIO-US B07 clinical trial (NCT00769470) in early-stage HER2-positive breast cancer received one cycle of neoadjuvant HER2-targeted therapy – including either trastuzumab, lapatinib, or both agents – followed by six cycles of the assigned HER2-targeted therapy plus docetaxel and carboplatin given every three weeks ^21, 22^. Core biopsies were obtained pre-treatment and on-treatment after 14-21 days of HER2-targeted therapy, and surgical excision specimens were obtained post-treatment (Figure 1a). We defined a discovery cohort of 28 patients for whom formalin-fixed paraffin-embedded (FFPE) samples were available from all three timepoints (pre-treatment, on-treatment, and at surgery). The cohort was balanced for both pCR and ER status (Extended Data Figure 1a-b) and was used for all exploratory analyses. We additionally defined an independent validation cohort of 29 patients from the same trial with matched pre- and on-treatment FFPE samples for evaluation of model performance.

**Figure 1.**
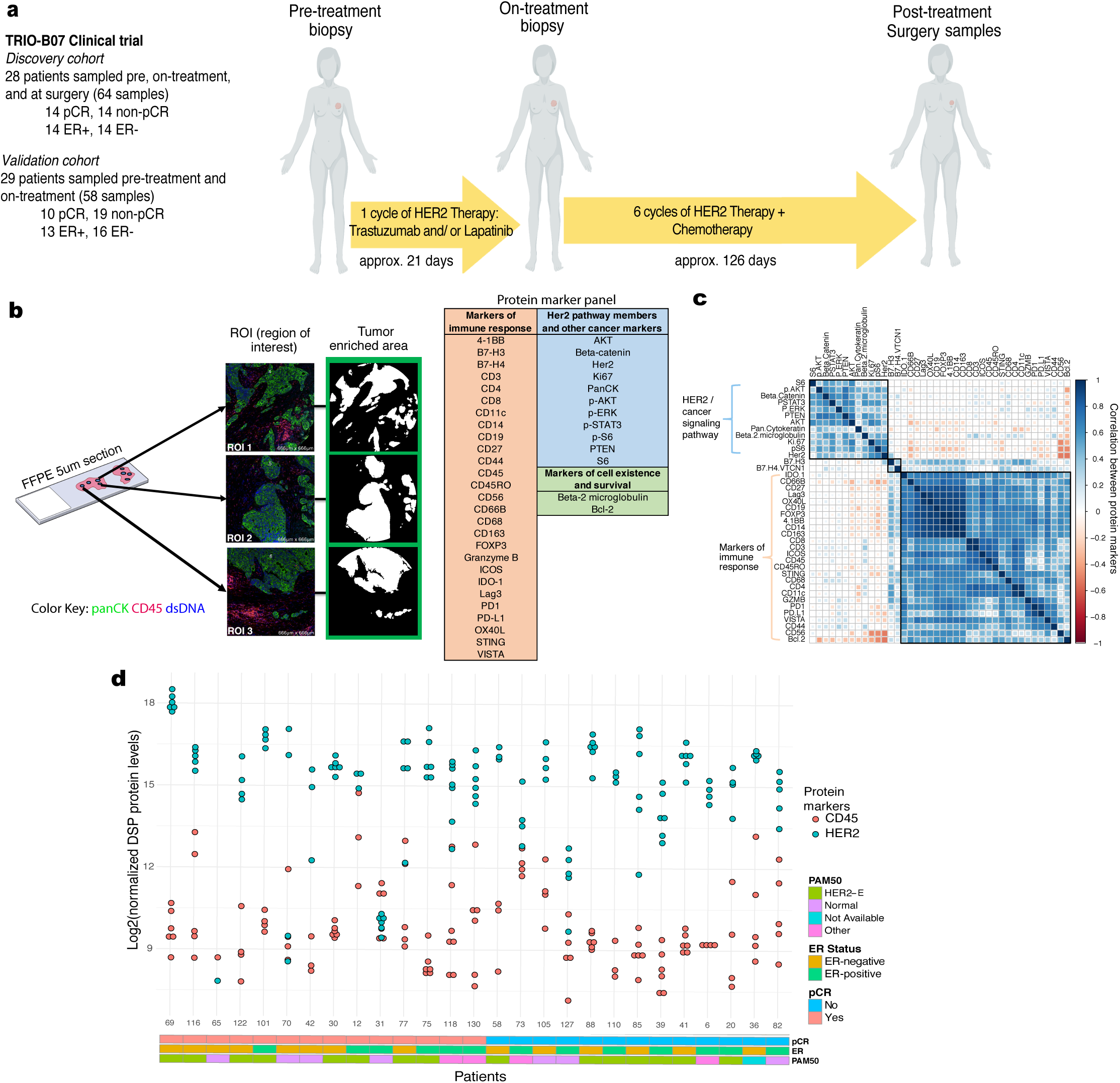
Spatial proteomic analysis of untreated HER2-positive breast tumors. a. Schematic overview of the discovery and validation cohorts analyzed with the GeoMx™ Digital Spatial Profiling (DSP) technology. Patients with invasive HER2-positive breast cancer enrolled on the TRIO-US B07 clinical trial were treated with one cycle of the assigned HER2-targeted therapy followed by six cycles of the assigned HER2-targeted treatment plus chemotherapy (docetaxel and carboplatin). Tissue was obtained at three timepoints (pre-treatment, on-treatment, and post-treatment/surgery). b. Multiple regions of interest (ROIs) per tissue sample were selected based on pancytokeratin enrichment (panCK-E) and subject to spatial proteomic profiling of 40 tumor and immune markers. Protein counts were measured within phenotypic regions corresponding to the PanCK-E masks that includes tumor cells and co-localized immune cells and separately for the inverted mask corresponding to panCK-negative regions. c. Pairwise correlation of pre-treatment protein marker expression across all ROIs in the discovery cohort. Black squares indicate probes in the same hierarchical cluster. d. Inter-tumor and intra-tumor variability in HER2 and CD45 protein expression in untreated HER2-positive breast tumors from the discovery cohort, where each point corresponds to an ROI. Clinical characteristics, including pCR status, estrogen receptor (ER) status, and PAM50 subtype (based on gene expression profiling) are indicated.

DSP enables multiplex proteomic profiling of FFPE tissue sections (Extended Data Figure 2a), where regions of interest (ROIs) can be selected based on both geographic and phenotypic characteristics (Extended Data Figure 2b). We employed a panCK enrichment strategy to profile cancer cells and colocalized immune cells across an average of four regions per tissue specimen (Figure 1b). Using CD45, panCK, and dsDNA immunofluorescent markers for visualization (Extended Data Figure 1c), we selected spatially separated regions, and a mask governing the UV illumination for protein quantitation was generated based on panCK immunofluorescence. In total, 40 tumor and immune proteins were profiled using DSP, and proteins assessed using both DSP and orthogonal technologies showed strong concordance (Figure 1b, Extended Data Figure 2c-e). We further utilized paired pre- and on-treatment bulk gene expression data from the same patients to infer PAM50 subtype and enable comparisons with the spatially resolved DSP data ^21, 22^.

**Figure 2.**
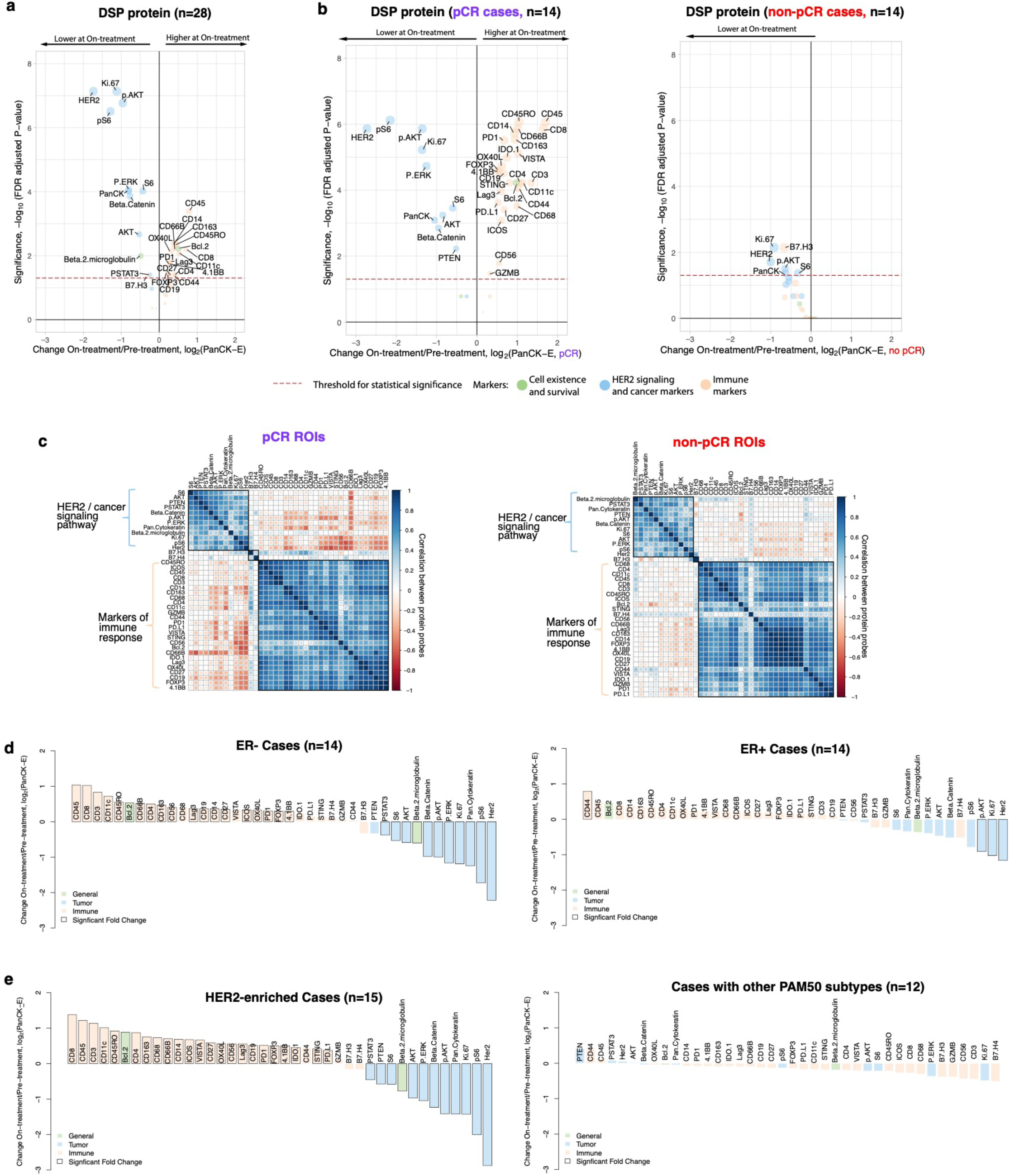
Spatial proteomic analysis reveals changes in cancer signaling and immune infiltration after short-term HER2 targeted therapy. a. Volcano plot demonstrating treatment-associated changes based on comparison of pre-treatment versus on-treatment protein marker expression levels in pancytokeratin-enriched (PanCK-E) regions. Significance, -log10(FDR adjusted p-value), is indicated along the y-axis. b. Volcano plots demonstrating treatment-associated changes in pCR versus non-pCR cases. c. Pairwise correlation of protein markers in pCR versus non-pCR cases. Black squares demarcate hierarchical clusters. d. Waterfall plots illustrating treatment-associated changes (pre-treatment to on-treatment) in ER+ and ER-cases based on protein expression. e. Waterfall plots illustrating treatment-associated changes in DSP protein expression (pre-treatment to on-treatment) in HER2-enriched and non-HER2-enriched cases (n=7 normal-like, n=2 luminal B, n=2 basal, n=1 luminal A). Analyses were performed in the discovery cohort.

In untreated tumors, the correlation amongst immune markers was striking, suggesting the coordinated action of multiple immune cell subpopulations (Figure 1c). HER2 pathway members and other downstream cancer signaling markers were also highly correlated, while the correlation between tumor and immune markers was minimal for most marker pairs. Inter- and intra-tumor variability at the proteomic level was evident prior to treatment, including for HER2 and the pan-leukocyte marker CD45 (Figure 1d). Averaging all ROIs per tumor to derive a composite score per marker, we found that tumors that achieved a pCR and those that did not had similar baseline HER2 levels (mean pCR cases: 14.50, mean non-pCR cases: 15.00) and baseline CD45 levels (mean pCR: 9.90, mean non-pCR: 9.64). Using a linear mixed-effects model with blocking by patient (Methods), we further found that individual DSP protein markers, including HER2 and CD45, did not significantly differ between pCR and non-pCR cases prior to treatment (unadjusted p > 0.10 for all markers).

### Reduced cancer signaling and increased immune infiltration after short-term HER2-targeted therapy

We used DSP to investigate treatment-related changes in breast tumor and immune markers during short-term HER2-targeted therapy by profiling on-treatment (after a single cycle of HER2-targeted therapy alone) biopsies in the discovery cohort. The protein markers that were most associated with pCR at the on-treatment timepoint were CD45 (unadjusted p=0.0024) and CD56, a natural killer (NK) cell marker (unadjusted p=0.0055) (Extended Data Figure 1d-e). We quantified the fold change in protein levels on-treatment relative to pre-treatment using a linear mixed-effects model with blocking by patient and visualized the significance (false discovery adjusted p-value) of all markers relative to their fold change in volcano plots. These analyses revealed a dramatic reduction in HER2 and Ki67, accompanied by other downstream pathway members, including pAKT, AKT, pERK, S6, and pS6, with the phosphorylated proteins decreasing comparatively more (Figure 2a). Immune markers – including CD45 and CD8, a marker of cytotoxic T-cells – exhibited the greatest increase in expression with treatment. Of note, increased expression of CD8+ T-cells was similarly observed in the TRIO-US B07 transcriptomic data through cell-type deconvolution ^21^. More generally, the on-versus pre-treatment bulk transcriptome data mirrored the changes seen at the protein level, but the fold changes were attenuated (Extended Data Figure 3). For example, using genes that correspond with the DSP protein markers, we found that the expression of HER2, AKT, Ki67, and breast cancer-associated keratin genes (KRT7, KRT18, and KRT19) ^26^ decreased significantly with treatment, while immune markers increased (Extended Data Figure 3). Despite the use of different analytes, measurements, and tissue sections, the DSP protein and bulk RNA datasets consistently showed reduced HER2 signaling and breast cancer-associated markers, accompanied by increased immune cell infiltration during neoadjuvant treatment. Given that lapatinib was associated with lower pCR rates in the TRIO-US B07 trial and more generally ^27^, we additionally assessed on-treatment changes in the trastuzumab-treated cases (arms 1 and 3, n=23) and observed similar patterns as in the full discovery cohort (Extended Data Figure 4a).

**Figure 3.**
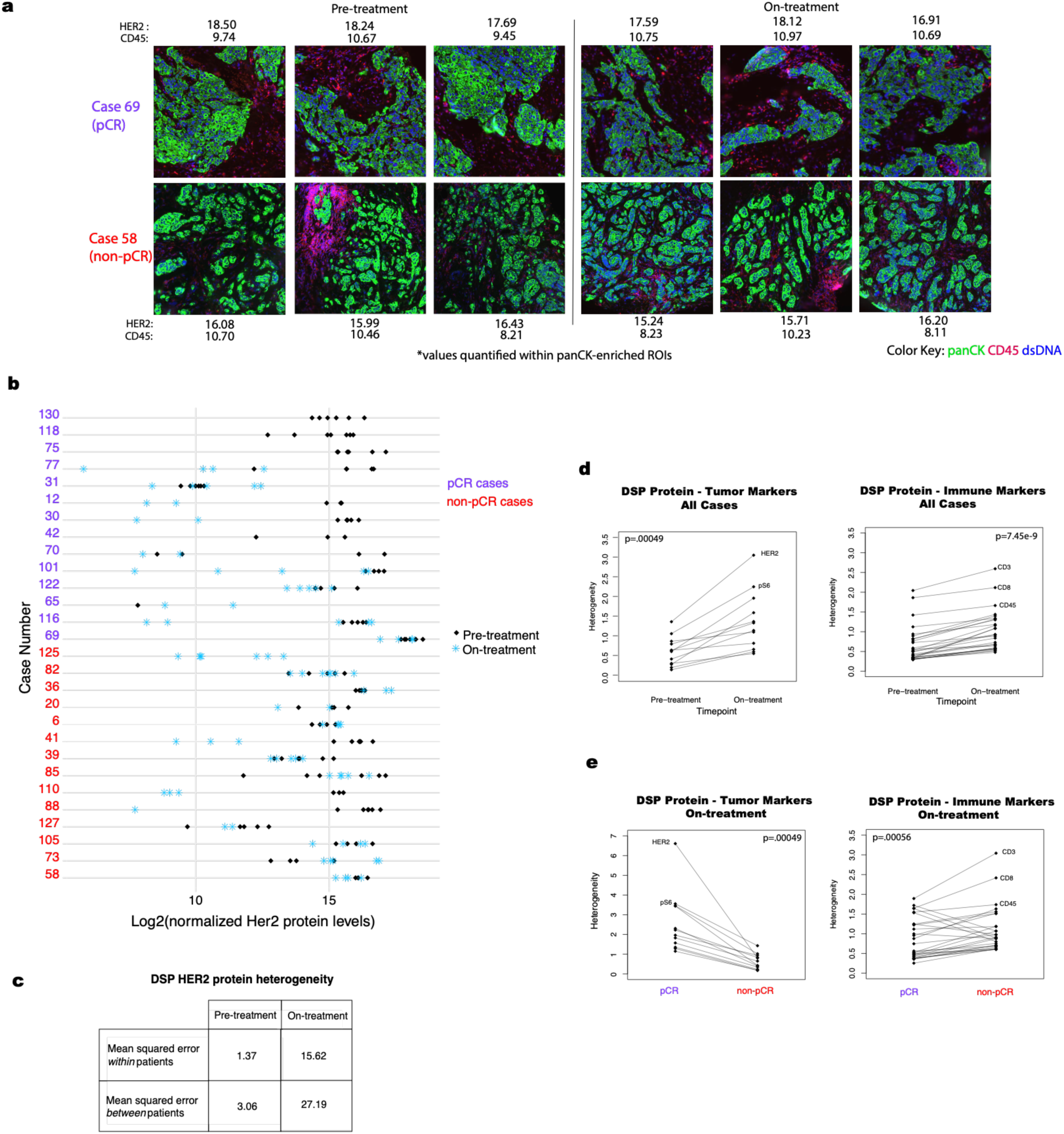
Increased heterogeneity of tumor and immune markers during HER2-targeted therapy. a. Representative images of ROIs from two cases and quantification of HER2 and CD45 protein levels (log2 normalized) in panCK-enriched regions. b. Comparison of DSP HER2 protein levels pre-treatment and on-treatment for all regions profiled per case per timepoint. c. Comparison of the mean squared error in DSP HER2 protein expression pre-treatment versus on-treatment within and between patients. d. Pre-treatment versus on-treatment heterogeneity for each DSP tumor and immune marker. e. On-treatment heterogeneity in DSP protein markers for pCR and non-pCR cases. Heterogeneity was calculated as the mean squared error within patients based on analysis of variance in panels c, d, and e. P-values are based on a two-sided paired Wilcoxon signed rank test. Analyses are based on the discovery cohort.

**Figure 4.**
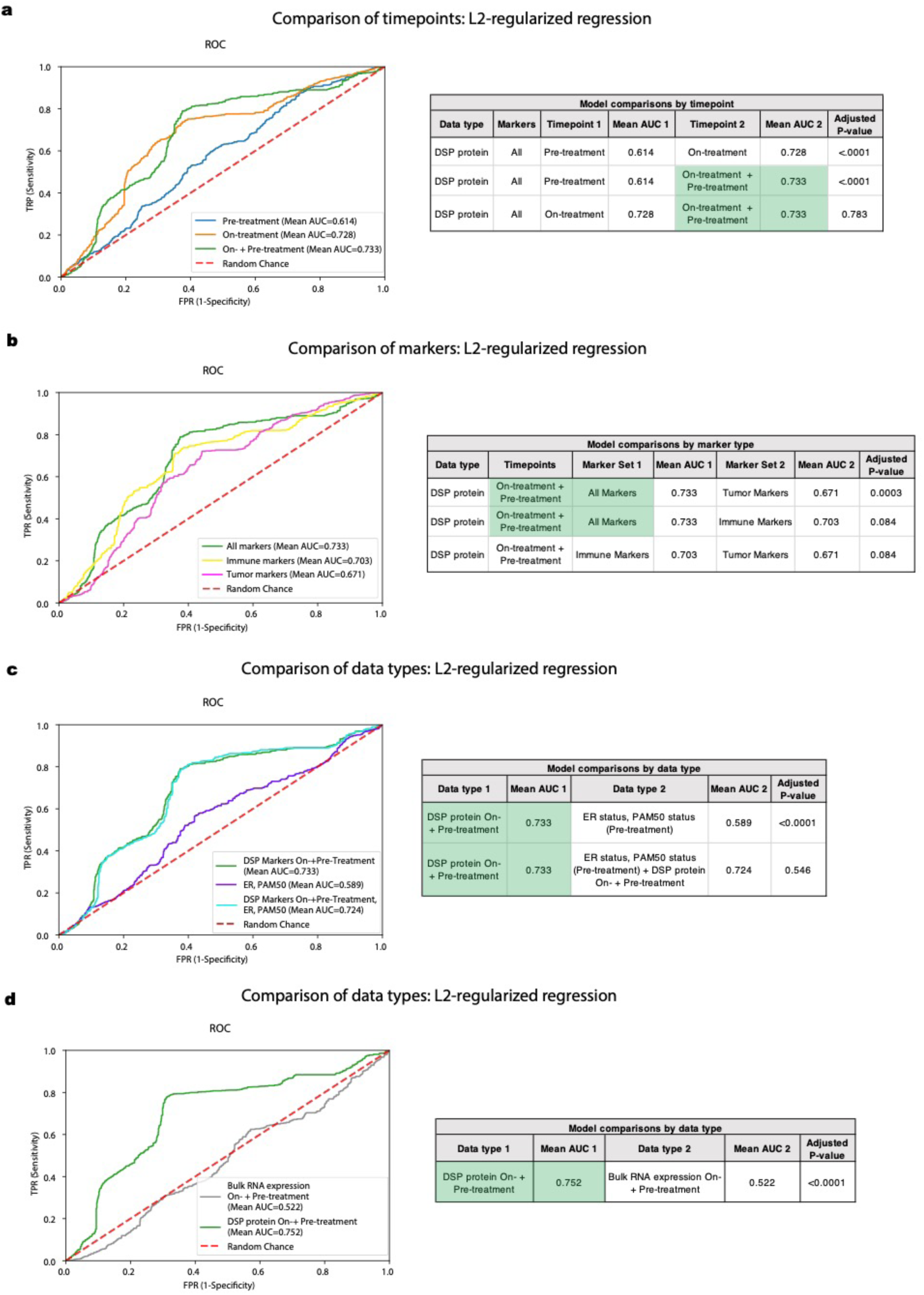
DSP of pancytokeratin-enriched paired pre- and on-treatment biopsies is associated with pathologic complete response in the discovery cohort and outperforms established markers. AUROC (Area Under Receiver Operating Characteristic) performance of various models were compared using nested cross-validation with Holm-Bonferroni correction for multiple hypotheses in the discovery (training) cohort. For (a-c), Receiver operating characteristic (ROC) curves were generated using cases with DSP panCK-enriched data from both the pre-treatment and on-treatment timepoints (n=23). a. ROC curves and statistical comparison of L2-regularized classifiers trained using DSP protein marker mean values (averaged across ROIs) pre-treatment, on-treatment and the combination of pre-treatment and on-treatment (“On-+ Pre-treatment”). b. ROC curves and statistical comparison of DSP protein On-plus Pre-treatment L2-regularized classifiers trained using all marker, tumor marker, and immune marker mean values. Cross-region mean marker values from both the pre-treatment and on-treatment timepoints were used in this analysis. c. ROC curves and statistical comparison of the On-plus Pre-treatment DSP protein L2-regularized classifier to a model trained using ER and PAM50 status. These two models were compared to a model that incorporates On-plus Pre-treatment DSP protein data, ER and PAM50 status. d. ROC and statistical comparison of On-plus Pre-treatment L2-regularized classifiers trained using DSP protein marker mean values versus bulk RNA expression using RNA transcripts corresponding to the DSP protein markers. ROC curves were generated using cases with DSP panCK-enriched data and bulk expression data from both the pre-treatment and on-treatment timepoints (n=21).

We next examined how treatment-associated changes differed based on tumor sensitivity to HER2-targeted therapy, stratifying tumors based on achievement of pCR following neoadjuvant therapy (Figure 2b). In the pCR cases, a multitude of immune markers increased with treatment, while, in the non-pCR cases, no significant treatment-associated immune changes were observed and the reduction in Ki67 and HER2 signaling was modest. These patterns can also be visualized via pairwise comparisons of protein marker correlations, which revealed a stronger negative correlation between the immune marker cluster and the cancer cell marker cluster in tumors that went on to achieve a pCR (mean fold change across all markers in pCR cases: −0.231, non-pCR cases: −0.075, two-sided Wilcoxon rank sum test p < 2.2e-16) (Figure 2c). Notably, no such differences between pCR and non-pCR cases were observed in the on-treatment bulk expression data from this same cohort ^21^.

Since both ER status ^8, 25^ and HER2-enriched subtype have been associated with response to neoadjuvant therapy ^10, 14, 18^, we explored how protein marker expression changed with these covariates. ER-negative tumors exhibited more significant changes on-treatment (relative to pre-treatment) compared to ER-positive tumors (mean absolute fold change ER-negative cases: 0.59, mean ER-positive cases: 0.36, two-sided Wilcoxon rank sum test p=0.0045, Figure 2d). However, when tumors were stratified by outcome, pCR cases exhibited more significant changes than non-pCR cases irrespective of ER status (Extended Data Figure 5a) and ER status was not predictive of pCR in this cohort (p=0.47). Similarly, tumors classified as HER2-enriched prior to treatment exhibited more significant changes in tumor and immune markers in the on-treatment biopsy relative to other subtypes (Figure 2e, Extended Data Figure 5b-c). For example, while CD8+ T-cells increased significantly with treatment in HER2-enriched cases, they decreased slightly in other cases. However, as in the full TRIO-US B07 transcriptomic cohort ^21, 22^, HER2-enriched subtype was not predictive of pCR (p=0.87).

**Figure 5.**
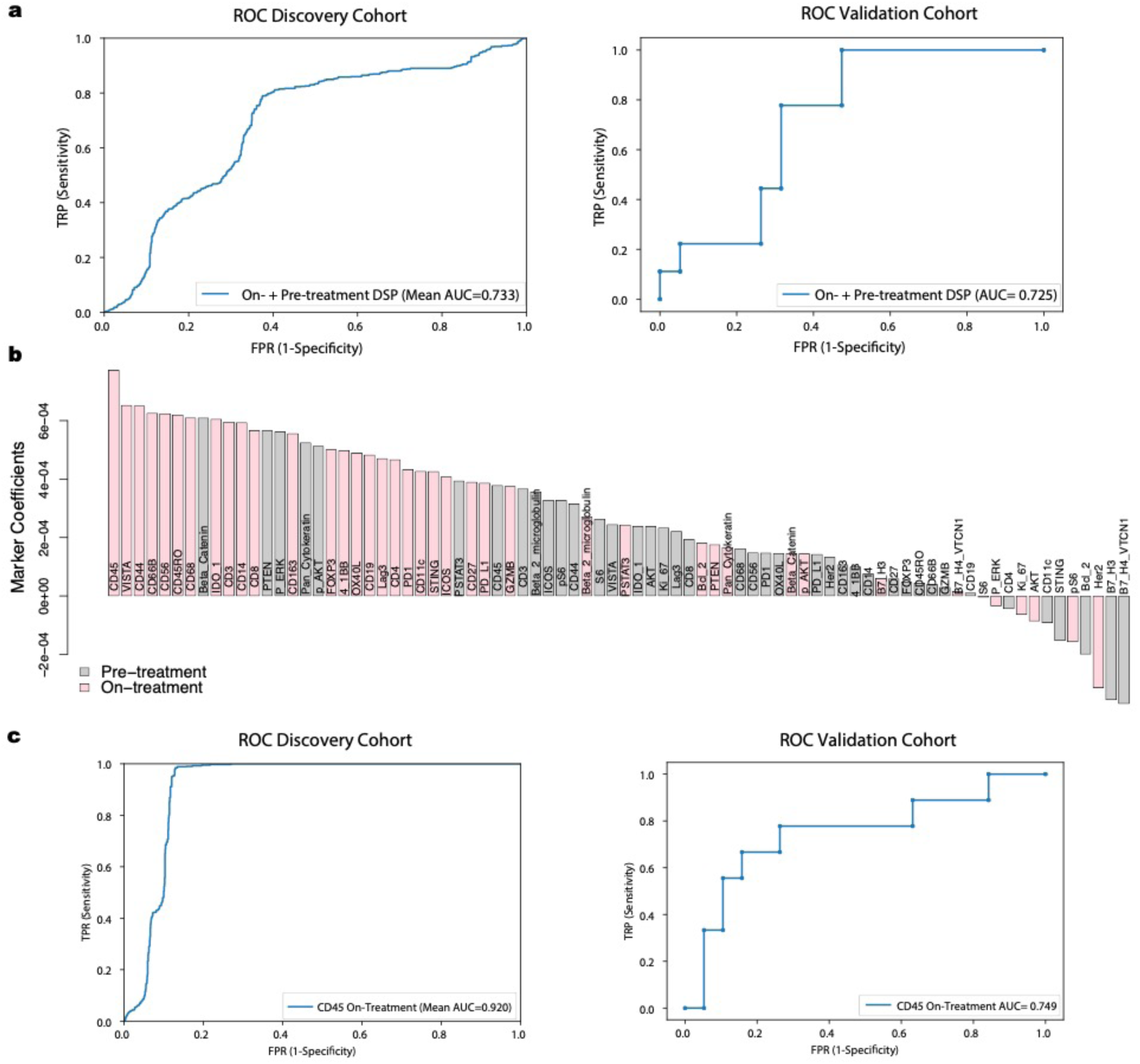
Validation of the DSP-based spatial proteomic biomarker in an independent cohort. a. Receiver operating characteristic (ROC) curves for On-plus Pre-treatment DSP protein L2-regularized classifier in the discovery (training) cohort (n=23, assessed via cross-validation) and the validation (test) cohort (n=28, assessed via train-test) using the 40-plex DSP protein marker panel. b. Coefficients for each of the 40 markers in the L2-regularized On-plus Pre-treatment DSP protein model, trained in the discovery cohort, and tested in the validation cohort. c. ROC curves for On-treatment L2-regularized classifier in the discovery (training) cohort (n=23, assessed via cross-validation) and the validation (test) cohort (n=28, assessed via train-test) using CD45 from the DSP protein marker panel.

In order to assess the utility of multi-region sampling, we measured changes on-versus pre-treatment using a single randomly selected region per tissue sample averaged across 100 simulations (Extended Data Figure 5d). Consistent with the findings based on all tumor regions, CD45 and CD8 showed the greatest increase on-treatment, while HER2 and pS6 decreased most in the single region analysis. While the magnitude of marker fold change with treatment was greater for pCR cases than non-pCR cases (mean absolute fold change across all markers in pCR cases: 0.87 versus non-pCR cases: 0.33, two-sided Wilcoxon rank sum test p= 1.02e-07), individual markers did not increase significantly with treatment in the single region analysis, reflecting increased variance.

We also examined treatment-associated changes in patients with residual tumor cells present at the time of surgery (non-pCR cases) to elucidate the biology associated with combined HER2-targeted therapy and chemotherapy. While the non-pCR cases showed limited changes at the on-treatment timepoint, by time of surgery there was a substantial reduction in the HER2 and downstream AKT signaling pathway, and a concomitant increase in immune markers in panCK-enriched regions (Extended Data Figure 1f). Notably, HER2 decreased more significantly than its downstream pathway members, which may reflect compensatory pathway activation contributing to resistance ^9, 28^. While some immune markers increased significantly in non-pCR cases at surgery (n=8), the fold change was diminished relative to pCR cases sampled on treatment (mean fold change non-pCR post-treatment: 0.30, mean fold change pCR on-treatment: 0.85, two-sided Wilcoxon rank sum test p=0.0021, Figure 2b). Amongst the immune markers that increased at surgery in the non-pCR cases, CD56 was most significant and potentially related to the role of NK cells in identifying and killing chemotherapy-stressed tumor cells ^29^. NK-cells were similarly found to increase at time of surgery in the TRIO-US B07 bulk expression data ^21, 22^.

### Increased heterogeneity of tumor and immune markers during HER2-targeted therapy

Given that genomic heterogeneity is a defining feature of HER2-positive breast cancer, we next evaluated the extent to which HER2 protein expression varied within different regions of a breast tumor biopsy through neoadjuvant treatment and between patients. As shown for two exemplary cases (Figure 3a), HER2 protein levels across geographically disparate regions within each tissue sample exhibited relatively consistent HER2 protein levels prior to treatment in the majority of cases (Figure 3b). Far greater heterogeneity in HER2 protein expression was observed on-treatment both between regions and between patients (Figure 3c). Similarly, CD45, a pan-leukocyte marker, showed increased heterogeneity between regions on-treatment and was higher in the pCR cases on-treatment as compared to the non-pCR cases (Extended Data Figure 6a). Such regional heterogeneity may reflect pharmacokinetic differences due to vasculature, tissue architecture, immune infiltration, or the biopsy itself, underscoring the importance of profiling multiple regions per sample on-treatment.

**Figure 6.**
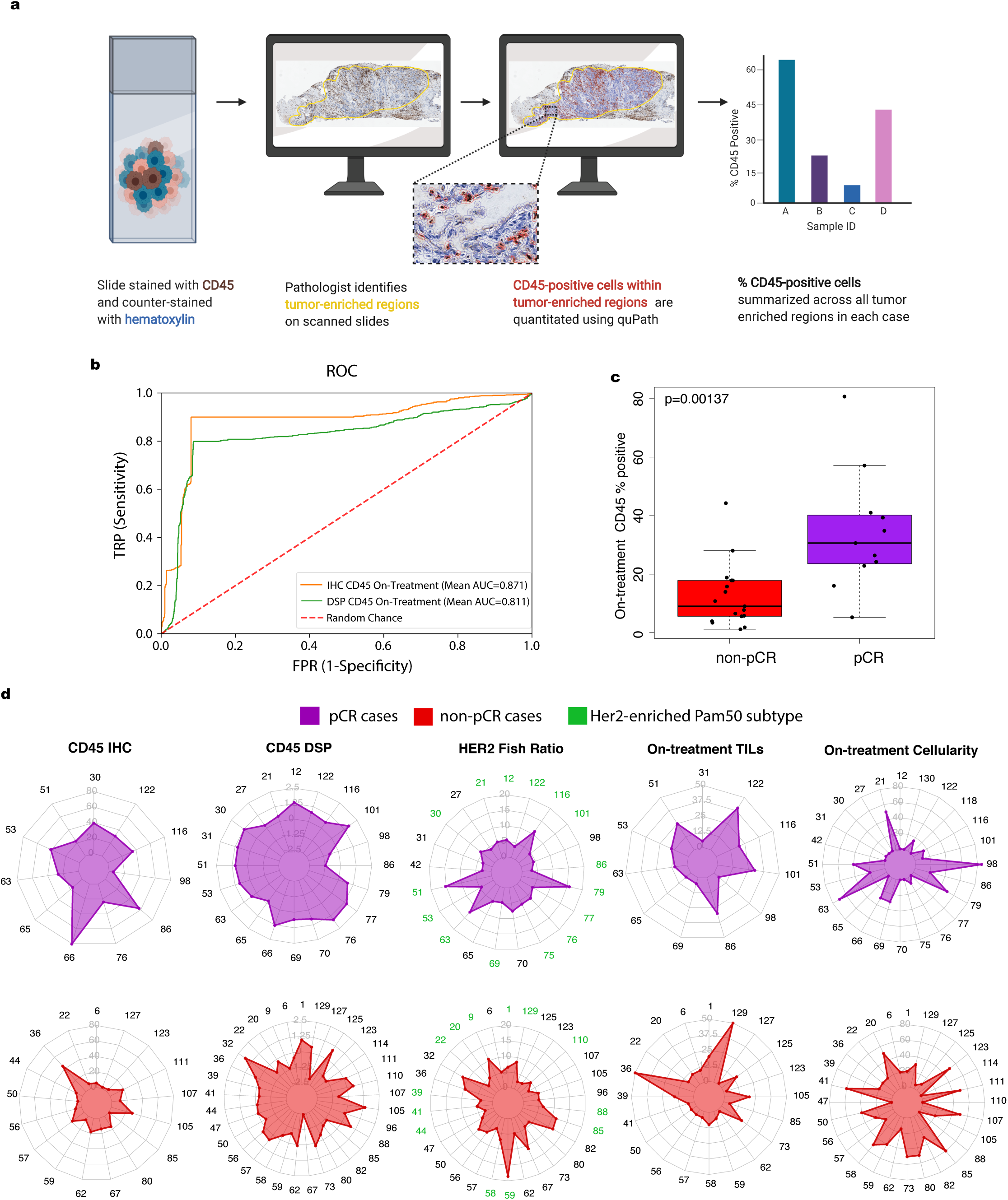
On-treatment IHC-based measurement of percentage CD45-positive cells in tumor-enriched regions is associated with pathologic complete response. a. Schematic showing process for quantitative assessment of CD45-positive cell counts from IHC. b. Receiver operating characteristic (ROC) curves for classifiers trained using either on-treatment DSP CD45 levels or on-treatment IHC-based CD45-positive percentage of cells. These models were evaluated on the subset of cases on which CD45 IHC was performed (n=28). c. Boxplot showing IHC CD45 % positive within tumor enriched regions stratified by pCR. P-values were derived using the Wilcoxon rank-sum test. For each boxplot, the colored box represents the interquartile range and the black lines extending from the box represent 1.5X the interquartile range. d. Radar plots showing on-treatment IHC CD45 % positive, on-treatment DSP CD45 levels, pre-treatment HER2 FISH ratios, on-treatment tumor infiltrating lymphocyte (TILs) score, and on-treatment tumor cellularity across patients. stratified by pCR status. All cases in which a given feature was measured across the DSP discovery and validation cohorts is shown.

We also investigated regional heterogeneity across all tumor and immune protein markers during treatment. For each marker and at each timepoint, regional heterogeneity across the cohort was computed as the within-patient mean squared error based on ANOVA (Methods). Across all markers, DSP protein heterogeneity increased significantly on-treatment relative to pre-treatment (Figure 3d), similar to that noted for HER2. These changes were widespread, with heterogeneity being higher for all tumor and immune markers on-treatment compared to pre-treatment. The probes with the greatest heterogeneity included both tumor (HER2, pS6) and immune (CD3, CD8) markers. Amongst tumors that failed to achieve a pCR, we evaluated heterogeneity throughout the course of neoadjuvant therapy. Heterogeneity amongst tumor markers was not significantly different on-treatment and pre-treatment (two-sided Wilcoxon rank sum test p=0.52), but increased at surgery (post-treatment), whereas immune marker heterogeneity increased on treatment with a subsequent decrease at surgery (Extended Data Figure 6b-c). Tumors that achieved a pCR exhibited higher protein heterogeneity amongst tumor markers (including HER2) on-treatment, whereas those that did not exhibited higher heterogeneity across immune markers (Figure 3e). Higher immune marker heterogeneity on-treatment in the non-pCR cases may reflect a less consistent immune response with some regions experiencing a greater immune influx than others. We did not observe higher pre-treatment HER2 heterogeneity in the non-pCR cases compared to pCR cases, as recently suggested based on FISH ^30^, and comparable regional heterogeneity amongst tumor markers was noted in pCR and non-pCR cases (Extended Data Figure 6b).

We further analyzed the DSP data to investigate the composition of immune cells in panCK-enriched regions (used in all other analyses) relative to the surrounding panCK-negative regions designed to capture the neighboring microenvironment (Extended Data Figure 7). Prior to treatment, both T cell (CD3, CD4, CD8) and macrophage (CD68) markers were more prevalent in the surrounding microenvironment, while CD56-positive NK cells and immunosuppressive markers (e.g. B7-H4 (VCTN1), PD-L1, IDO) were higher in panCK-enriched regions (Extended Data Figure 7b). Although we cannot rule out the possibility of non-specific staining, these findings are consistent with T cell exclusion, where IDO and PD-L1 are thought to impair intratumor proliferation of effector T cells ^31^. A similar immune profile was observed during HER2-targeted therapy alone. However, post-treatment, in the non-pCR cases with tumor remaining, most immune markers were more prevalent in the panCK-enriched regions, compared to the neighboring microenvironment, including CD8 and CD68 (Extended Data Figure 7b). Both prior to treatment and on-treatment, immune cell localization was similar in patients that achieved a pCR and those that did not (Extended Data Figure 7c), as well as for ER-positive versus ER-negative cases (Extended Data Figure 7d).

As proof of principle that other multiplexed imaging technologies can similarly be used to profile panCK-enriched tumor, we also used multiplex immunohistochemistry (mIHC) to profile tissue samples from a patient that achieved a pCR and one that did not. panCK antibodies were to define mask regions and several markers that changed significantly with treatment based on DSP – namely HER2, CD45, and CD8 – were quantified across the whole tissue section and within panCK-enriched regions (Extended Data Figure 8a-b). As expected, changes in protein expression signals were muted when the whole tissue section was considered relative to panCK-enriched regions. These data further support the concept that panCK enrichment may be beneficial for defining tumor and co-localized immune changes in breast and other tumors.

The geospatial distribution of tumor and immune cells has been associated with relapse and survival in multiple tumor types ^32, 33^. We therefore investigated the relationship between treatment and the tumor-microenvironment border using perimetric complexity, which is proportional to the perimeter of a region squared, divided by the area of the region ^34^ (Methods, Extended Data Figure 8c). Prior to treatment, no significant difference in perimetric complexity was observed between pCR and non-pCR cases (p=0.299, Extended Data Figure 8d). However, perimetric complexity decreased significantly on-treatment relative to pre-treatment (p=1.32e-6, Extended Data Figure 8e). These data suggest that treatment may affect the geographic distribution of tumor cells as well as tumor cell content. Indeed, the proliferative marker, Ki67, was highly correlated with perimetric complexity (Extended Data Figure 8f). Thus, for highly proliferative tumors, the perimeter of the tumor-microenvironment border may be relatively larger, allowing for increased crosstalk with the surrounding microenvironment.

### DSP of paired pre- and on-treatment biopsies reveals features associated with pCR

Given the dramatic differences in treatment-associated changes in pCR cases compared to non-pCR cases (Figure 2b), we next sought to evaluate whether DSP protein marker status prior to treatment or early during the course of therapy could be used to predict pCR. We used an L2-regularized logistic regression to classify tumors by pCR status based on average DSP protein expression levels across multiple ROIs profiled pre-treatment, on-treatment, or the average marker expression both pre-treatment and on-treatment (denoted “on-plus pre-treatment”) and evaluated model performance via nested cross validation within the discovery cohort (Methods). Cases with tumor-enriched regions identified via DSP at both timepoints were utilized in this analysis (n=23 cases, Extended Data Figure 1b). A model based on on-treatment protein expression outperformed that based on pre-treatment protein expression (mean AUROC=0.728 versus 0.614) and performed comparably to a model incorporating both on-treatment and pre-treatment protein expression levels (mean AUROC=0.733) (Figure 4a). A classifier trained using both immune and tumor markers outperformed a model using tumor markers alone (Figure 4b).

For the DSP protein on-plus pre-treatment classifier, we investigated the importance of multi-region sampling and heterogeneity by extending the model to incorporate both the mean marker expression across all regions and the standard error of the mean (SEM) for each marker between regions (Methods). This analysis was restricted to patients with at least 3 regions profiled at both timepoints (n=16, Methods). We found that utilizing the mean immune values and the SEM for tumor markers outperformed a model based on mean values for both tumor and immune markers (Extended Data Figure 9a), suggesting value in capturing heterogeneity amongst tumor markers.

We next compared the performance of the DSP protein on-plus pre-treatment classifier with features previously associated with outcome (ER status and PAM50 subtype), where models were again evaluated via cross-validation in the discovery cohort. Of note, a model based on ER status and HER2-enriched PAM50 status performed poorly in this cohort (mean AUROC=0.589) and the addition of these two features or additional pathologic features to the DSP protein on-plus pre-treatment data set did not improve the AUROC (Figure 4c, Extended Data Figure 9b-d). Given the availability of bulk transcriptomic data for these cases, we also evaluated a model using paired on- and pre-treatment bulk RNA expression data for the 37 markers that overlapped with the DSP protein panel. This model also performed significantly worse than that based on the DSP protein data (Figure 4d, p<0.0001 via cross-validation). This is not surprising since amongst the 37 overlapping DSP and bulk RNA expression markers, only 16 were positively correlated pre-treatment (Extended Data Figure 3b-c). Various factors may contribute to the lack of strong correlation between protein and RNA expression levels, including panCK enrichment, RNA transience/degradation ^35^, and post-translational regulation ^36^, where protein expression is a more proximal readout of cellular phenotype.

### DSP predicts pCR in an independent validation cohort

In light of these promising findings, we further sought to evaluate the performance of the DSP protein on-plus pre-treatment classifier in an independent cohort (n=29) of patients from the TRIO B07 clinical trial (Extended Data Figure 9e). As with the discovery cohort, an average of four panCK-positive regions were profiled from each pre- and on-treatment tumor tissue and the same panel of 40 protein antibodies was utilized. The change in markers on-treatment relative to pre-treatment mirrored that observed in the discovery cohort (Extended Data Figure 9f): T-cell markers (CD3, CD4, and CD8) increased while the tumor markers HER2 and Ki67 showed the most significant reduction. Similar to the discovery cohort, in the validation cohort, we observed that on-treatment versus pre-treatment protein expression differences were more dramatic in tumors that ultimately underwent pCR (Extended Data Figure 9g). AUROC performance of the L2-regularized logistic regression model, trained in the discovery cohort, was evaluated in the validation (test) cohort. The performance of the DSP protein on-plus pre-treatment model in predicting pCR was comparably high in the discovery (assessed via cross-validation, mean AUROC = 0.733) and validation (assessed via train-test, AUROC=0.725) cohorts (Figure 5a). In the on-plus pre-treatment classifier, which was trained in the discovery cohort and tested in the validation cohort, the marker with the largest L2-regularized coefficient was on-treatment CD45 protein levels. Other features with large coefficients included on-treatment markers that represent tumor-infiltrating lymphocyte and macrophage populations (CD44, CD66B) (Figure 5b). On-treatment HER2 protein expression had a negative coefficient in the model, consistent with poor outcome being associated with high HER2 levels during treatment.

In light of the widespread use of trastuzumab in current neoadjuvant treatment paradigms ^37^, we further assessed model performance for the best performing on-plus pre-treatment DSP protein model in the trastuzumab-containing cases (arms 1 and 3, n=19). Again, similar model performance and marker coefficients were seen as in the full discovery cohort and in the subset of cases in the validation cohort who were treated with trastuzumab (Extended Data Figure 4b-d). The validation of these findings in an independent cohort demonstrates the robustness of this approach and potential utility of multiplex spatial proteomic profiling to identify novel candidate biomarkers.

Given that CD45 on-treatment values had the greatest coefficient in the model (Figure 5b), we assessed model performance using this single marker by training an L2-regularized logistic regression model in the discovery cohort and evaluating this model in the validation (test) cohort. The performance of the DSP CD45 on-treatment model was high in both the discovery (assessed via cross-validation, mean AUROC=0.920) and validation (assessed via train-test, AUROC=0.749) cohorts (Figure 5c).

### On-treatment CD45-positive cell percentage in tumor-enriched regions is associated with pathologic complete response

Building on the striking finding that a single immune marker (CD45) predicted pCR with high accuracy, we sought to investigate whether quantitative DSP measurements correlated with conventional IHC (Figure 6a). Using 28 cases from a combination of the discovery and validation cohorts with additional tissue remaining (Extended Data Figure 10a) we performed CD45 IHC and quantified the percentage of CD45 positive cells within tumor-enriched regions from the on-treatment timepoint (Methods). PanCK-enriched CD45 measurements from DSP correlated with the CD45 IHC measurements (r=0.53) (Extended Data Figure 10b), as well as with stromal tumor-infiltrating lymphocytes (TILs) (r=0.41). Using the sub-cohort with CD45 IHC data, performance was strong in both a model trained using on-treatment IHC CD45-positive cell percentage (mean AUROC=0.871) and a model trained using on-treatment DSP CD45 (mean AUROC=0.811, both models assessed using cross-validation, Figure 6b). Cases that ultimately achieved a pCR tended to have higher CD45 staining (Figure 6c) and higher CD45 DSP levels (Figure 6d, Extended Data Figure 10c) in tumor-enriched areas on treatment, and this separation between pCR and non-pCR cases was greater for both CD45 measurements than for on-treatment stromal TILs, on-treatment tumor cellularity, or pre-treatment HER2 FISH ratios (Figure 6d, Extended Data Figure 10c). In light of the significant separation between pCR and non-pCR cases using CD45 IHC and CD45 DSP levels, we next used each feature to establish thresholds to predict pCR; in the case of CD45 IHC, the selected threshold was 20% CD45-positive cells (Extended Data Figure 10d). Positive predictive value (PPV) is among the most relevant model performance parameters in early neoadjuvant treatment selection, where the goal is to de-escalate treatment for tumors that are highly likely to undergo pCR while not depriving others of potentially curative therapy: in the CD45 DSP validation set, the PPV was 0.714, modestly lower than the CD45 DSP discovery set (0.750) and the CD45 IHC discovery set (0.818) (no validation set was available for CD45 IHC). Performance was similar when excluding the lapatinib-only arm (Extended Data Figure 4e). Overall, these findings demonstrate the potential utility of profiling a single marker in an on-treatment biopsy to predict which patients will respond early during HER2-targeted therapy, such that subsequent therapy could be tailored accordingly.

## Discussion

Bulk genomic and transcriptomic profiling has been a mainstay of cancer biomarker discovery efforts in recent years. However, admixture amongst heterogeneous cellular populations complicates the analysis of such data, issues which are compounded when studying longitudinal samples, where the changing composition and localization of cell populations may reflect the biology of disease progression or mechanisms of treatment response. Indeed, efforts to establish validated biomarkers of response to HER2-targeted therapy based on bulk genomic and transcriptomic profiling have met with limited success to date in other trial cohorts and in TRIO-US B07 ^10, 12, 21^. We reasoned that *in situ* proteomic profiling of the tumor-immune microenvironment through therapy would circumvent the limitations of dissociative techniques and improve our ability to uncover features associated with response to neoadjuvant HER2-targeted therapy. Here we used the DSP technology ^24^ to simultaneously profile 40 tumor and immune markers *en bloc* on a single 5μm section of archival tissue from breast tumors sampled before, during, and after neoadjuvant HER2-targeted therapy in the TRIO-US B07 clinical trial. In order to enhance signal while accounting for intra-tumor heterogeneity, we employed a pan-CK masking strategy to enrich for tumor cells and co-localized immune cells across multiple regions, each comprised of ∼300-600 cells, per sample. Our results illustrate the feasibility and power of multiplex *in situ* proteomic analysis of archival tissue samples to provide proximal readouts of tumor and immune cell signaling through therapy. Many signaling proteins/phospho-proteins, including those profiled here, are considered protein network bottlenecks and integrate mutational and transcriptional changes ^38, 39^, making this a particularly powerful approach to characterize compensatory signaling and mechanisms of resistance.

DSP of longitudinal breast biopsies from this trial cohort uncovered changes associated with therapy, including markedly decreased HER2 and downstream AKT signaling on-treatment, accompanied by increased CD45 and CD8 expression, consistent with infiltrating leukocytes and cytotoxic T-cells, respectively. By the time of surgery, following a full course of neoadjuvant therapy, the tumor-immune composition changed considerably with increased CD56 expression in non-pCR cases, potentially reflecting NK cell-mediated killing of chemotherapy-stressed tumor cells ^29^. While immune influx on-treatment was also observed in the bulk expression data from this cohort ^21^, there it did not correlate with response, leaving open the question of whether the biopsy itself may have induced these changes. DSP, in contrast, revealed that changes in both tumor and immune markers on-treatment were more dramatic in tumors that went on to achieve a pCR and, critically, on-treatment protein expression robustly predicted response in an independent validation cohort. Neither pre-treatment protein expression, bulk pre- and on-treatment gene expression data, nor established predictive features were predictive in this cohort, emphasizing the power of this quantitative multiplexed spatial proteomic biomarker. This work also illuminates study design considerations, including the value of panCK enrichment of tumor cells (or other markers to enrich for specific cell populations) and multi-region profiling to capture regional tumor heterogeneity and treatment-associated changes, concepts that should be broadly applicable to biomarker discovery in other epithelial tumor types. Our findings thus address a critical unmet clinical need given the considerable emphasis devoted to identifying subsets of the population in which therapy should be escalated, for example by combining HER2-targeted agents, or safely de-escalated, for example through shortening or omission of chemotherapy and its associated toxicities ^40, 41^. While numerous biomarkers have been considered to help guide personalized targeting of escalated versus de-escalated approaches in early-stage HER2-positive breast cancer – including imaging, circulating tumor DNA, and pre-treatment immune scores or intrinsic subtype – there is currently no validated biomarker that can guide patient stratification. The increasing plethora of options for HER2-targeted therapy, including novel highly effective but potentially toxic agents ^42, 43^, combined with great heterogeneity in response make HER-positive+ breast cancer the ideal setting for the development of optimally personalized therapy over the next decade. The next step towards clinical translation of the novel biomarker described here based on CD45 protein expression or cell counts will involve prospective validation in a neoadjuvant clinical trial that selects therapy based on biomarker status and assesses both pCR and RFS ^44^.

## Online Methods

### Cohort selection

The TRIO-US B07 clinical trial was a randomized, multicenter study that included 130 women with stage I-III unilateral, HER2-positive breast cancer^21, 22^. The IRB at the University of California Los Angeles (UCLA) approved the clinical trial TRIO-US B07 (08-10-035). The IRB at Stanford approved the use of the TRIO-US B07 clinical trial specimens for correlative studies in the Curtis Lab (eProtocol #32180). Informed consent was obtained from all participants. This covers consent from patients for their samples to be shared with other researchers. Enrolled patients were randomly assigned to three treatment groups, dictating the type of targeted therapy namely trastuzumab, lapatinib, or trastuzumab and lapatinib in combination. Breast tumor biopsies were obtained prior to treatment and following 14-21 days of the assigned HER2-targeted therapy (without chemotherapy), which was followed by six cycles of the assigned HER2-targeted treatment plus docetaxel and carboplatin given every three weeks and surgery. For each timepoint, core biopsies or surgical tissue sections were obtained and stored as either fresh frozen or FFPE material. In total, 28 cases with FFPE samples available from all three timepoints (pre-treatment, on-treatment, and at surgery) were selected for inclusion in the discovery cohort based on sample availability and quality, with balancing by pCR status and ER status (Figure 1a, Extended Data Figure 1a). An additional 29 cases with FFPE samples available pre-treatment and on-treatment were selected for the validation cohort in order to assess performance of the classifier (Extended Data Figure 9e). Of note, the validation cohort was used exclusively to evaluate model performance. All other analyses are based on the discovery cohort. Tumor cellularity was assessed by a subpecialty breast pathologist (GRB) using tumor sections stained with hematoxylin and eosin. Samples with cellularity of 0 were omitted from further analysis. For other tissue sections estimated to have a cellularity of 0, tumor cells were identified on the FFPE sections used to perform DSP (distinct from the H&E sections used for pathology review), and these were included in the analysis (Extended Data Figure 1b).

### DSP Data Generation and Analysis

Digital Spatial Profiling (DSP, NanoString) was performed as previously described ^24^. In brief, tissue slides were stained with a multiplexed panel of protein antibodies contained a photocleavable indexing oligo, enabling subsequent readouts (Extended Data Figure 2a). Regions of interest (ROIs) were selected on a DSP prototype instrument and illuminated using UV light. Released indexing oligos from each ROI were collected and deposited into designated wells on a microtiter plate, allowing for well indexing of each ROI during nCounter readout (direct protein hybridization). Custom masks were generated using an ImageJ pipeline, as described previously ^45^. For each tissue sample, counts for each marker were obtained from an average of four (range 1-7) panCK-enriched (panCK-E) ROIs. Raw protein counts for each marker in each ROI were generated using nCounter ^46^. The raw counts were ERCC-normalized (based on the geometric mean of the three positive control markers). Histone H3 was used as a housekeeping marker and ROIs with extreme Histone H3 (more than three standard deviations away from the mean) were filtered (< 1% of ROIs). The geometric mean of two IgG antibodies were used to calculate the background noise and we noted markers with signal to noise ratio <3x (Extended Data Figure 2f). Immune markers were normalized based on ROI area to measure total density of immune content in the region. Tumor markers were normalized using the housekeeping antibody (Histone H3) in order to capture the status of the cancer signaling pathways on a per cell basis. As further quality control, area normalization factors and housekeeping normalization factors were compared per ROI, and ROIs were filtered with disparate normalization factors across the two methods (this represented 6% of all ROIs). All normalized counts were converted to log2 space for downstream analysis. The analyses carried out in this study are comparative in nature (e.g. pre-treatment vs on-treatment, pCR vs non-pCR) and are robust to variations in normalization methods.

### Bulk mRNA Expression Analysis

Bulk mRNA Expression Analysis was performed as described elsewhere^21^. In brief, RNA was extracted using the RNeasy Mini Kit (Qiagen), quantified by the Nanodrop One Spectrophotometer (ThermoFisher Scientific). RNA samples were labeled with cyanine 5-CTP or cyanine 3-CTP (Perkin Elmer) using the Quick AMP Labeling Kit (Agilent Technologies). Gene Expression Microarray experiments were performed by comparing each baseline sample to samples taken after 14-21 days of HER2-targeted therapy (on-treatment). Each on-treatment sample was compared to the pre-treatment sample from the same patient. Limma^47, 48^ was used for background correction (“normexp”), within-array normalization (“loess”), between-array normalization, and for averaging over replicate probes. For the downstream analyses, including batch correction and comparisons with the DSP cohort, the normalized counts were converted to log2 space. Combat ^49^ was used to remove potential batch affects associated with microarray run date. PAM50 status pre-treatment and on-treatment was inferred using AIMS (Absolute Intrinsic Molecular Subtyping), an N-of-1 algorithm that is robust to variations in data set composition ^50^. This approach was utilized given the expected preponderance of HER2-enriched cases in this cohort.

### Correlation Analyses

Plots showing the correlation between protein markers (Figure 1c, 2c) were overlapped with hierarchical clustering in the form of black squares. The difference between the distribution of correlation values in pCR versus the non-pCR cases was evaluated using a two-sided Wilcoxon two-sample t-test. For the correlation between DSP protein data and bulk RNA data, the Spearman rank correlation was computed for pre-treatment samples using the average of all DSP ROIs (both panCK-enriched ROIs and surrounding microenvironment-enriched ROIs) per patient. For the correlation between DSP protein data and bulk RNA data, the Spearman rank correlation was computed per region. For all other correlation analyses (Extended Data Figure 2c,e; Extended Data Figure 10b) Pearson correlation was computed per tissue.

### Comparative Analyses

For the comparative analyses (e.g. pre-treatment vs on-treatment, pCR vs non-pCR, panCK-enriched vs panCK-negative) of DSP protein data, where multiple regions were sampled per patient, we utilized a linear mixed-effects model with blocking by patient ^51^. This model allows for marker levels to be compared in a patient-matched manner while controlling for differences in the number of ROIs profiled per patient. The coefficient of the fixed effect is the change attributable to that variable (x-axis of volcano plots), and the p-value used to calculate false discovery rates (y-axis of volcano plots) is based on the t-value (a measure of the size of the difference relative to the variation in the sample data). False discovery rates (FDR) were computed using the Benjamini & Hochberg procedure ^52^, and an FDR-adjusted p-value of 0.05 was set as the significance threshold.

### Region Subsampling

We sought to investigate the impact of utilizing a single randomly selected region per tissue sample, rather than multiple regions, when assessing on-versus pre-treatment protein expression changes. For these analyses (Extended Data Figure 5d), we performed 100 iterations in which a single region was selected from each tissue and computed fold changes and corresponding p-values averaged over these 100 experiments. The number of random samplings was chosen empirically by raising the number of iterations beyond the number required to make the resulting output robust to further increases in the number of iterations used (p-value convergence).

### L2-regularized Logistic Regression Using Molecular Data

#### Models and features

L2-logistic regression using liblinear as a solver was used for classification of pCR vs non-pCR cases. DSP marker values pre-treatment and on-treatment were averaged across all ROIs to derive a composite value for each marker for that timepoint. Five patients were excluded from the DSP models because data was available only at a single timepoint (Extended Data Figure 1b). Mean DSP marker expression features were used in models comparing patient timepoints, tumor versus immune markers, DSP protein features versus established predictive features (ER status and PAM50 classification), and DSP protein versus Bulk RNA features (using RNA gene transcripts corresponding to DSP protein markers). To assess heterogeneity, standard error of the mean (SEM) was calculated for marker values across all ROIs for tissues with at least 3 ROI to derive a composite value for each marker for that timepoint. These SEM features were used in combination with mean expression features in models assessing the predictive value of heterogeneity. Additional single feature models were compared in a sub-cohort with IHC data available (Extended Data Figure 10a): CD45 DSP on-treatment with CD45 IHC on-treatment. Generation of CD45 IHC data is described below.

#### Model comparisons and evaluation of performance via internal cross-validation

Model performance was evaluated and models compared using nested cross-validation using the python package sklearn ^53^. Data were divided into N folds using stratified sampling (“stratified cross-validation”). The number of folds was chosen based on the number of cases in the non-pCR group (the class with fewer cases) such that the testing data would contain two cases from each class. Each model was trained using N-1 folds and scored using mean AUROC on the remaining fold. This process was iteratively repeated holding out a different fold each time. The L2-penalization weight was chosen using stratified cross-validation within the N-1 training dataset, with the weight associated with highest mean accuracy within this inner cross-validation selected for scoring. This nested cross-validation process was repeated 100 times using randomly generated folds. Model scores were then compared using an unpaired two-sided t-test with Holm-Bonferroni correction for multiple hypotheses. ROC curves were generated by averaging across the ROC curves from the 100 repeats of N-fold cross-validation, with each repeat containing a different random split of folds.

#### Evaluation of model performance in an independent validation cohort

As described above, marker values pre-treatment and on-treatment were averaged across all ROIs to derive a composite value for each marker for that timepoint. Model selection was carried out using cross-validation as described above. The best performing model was selected and trained using the entire discovery cohort. Finally, model performance based on the AUROC was evaluated in the independent validation (test) cohort. For the models assessing CD45 IHC, cross-validation was used, as described above, to assess model performance.

### Metrics of Heterogeneity

Marker heterogeneity was calculated as the mean squared error from the analysis of variance done on a linear model with marker values as the dependent variable and patient identity as the independent variable (the data set was subsetted to the particular timepoint or clinical outcome of interest).

### Perimetric Complexity

Perimetric complexity was computed for the panCK-enriched binary masks for each ROI using ImageJ^34^. A linear mixed-effects model^51^ with blocking by patient was used to the compare the perimetric complexities of all the panCK-enriched regions pre-treatment and on-treatment regions and for cases that achieved a pCR versus those for cases that did not achieve a pCR.

### Multiplex IHC Analysis

Unstained, paraffin-embedded sections were analyzed by multiplex IHC analysis used the following markers: PanCK (AE1/AE2), CD8, CD45 LCA, and HER2 (29D8 CST). Stained samples were scanned, digitized as a series of square sub-images (“stamps”), and visualized using HALO. PanCK masking and tissue area masking was performed on each stamped tissue region using Fiji (ImageJ)^54^. Briefly, the PanCK channel was used to generate the masks (using the following ImageJ tools: Enhance Contrast, Threshold, Dilate, Fill Holes, Create Selection) for the panCK-positive region and the entire tissue region and CD8, CD45, and HER2 were quantified within each masked region (using the ImageJ Measure tool). A weighted average (with weights corresponding to each mask area) was used to calculate CD8, CD45, and HER2 levels across all the scanned sub-images that comprise the tissue (either tissue mask or panCK mask area).

### CD45 IHC Analysis

The concordance between CD45 DSP protein levels and CD45 counts based on immunohistochemical (IHC) staining and digital quantification was evaluated in 28 on-treatment cases with remaining tissue. IHC was performed on the Ventana BenchMark Ultra instrument (Ventana Medical Systems, Tucson, AZ) according to manufacturer’s protocols using primary antibody specific for CD45 (clone 2B11+PD7/26, Ventana, predilute) and CC1 antigen retrieval. Slides were counter-stained with Hematoxylin, and then scanned by Leica Aperio ScanScope systems. An adjacent 5µm paraffin sections were also cut for standard H&E staining. A board-certified pathologist specializing in breast cancer (GRB) reviewed each corresponding H&E slide and circled the tumor-enriched regions on CD45-stained images, which were then analyzed using QuPath image analysis software (v.0.2.0-m9)^55^. Pathologist-circled tumor-enriched regions were used for downstream analysis to mirror the pan-CK enrichment strategy employed in the DSP studies. Cells positive for CD45 were identified as having mean cell 3, 3’-diaminobenzidine (DAB) OD mean (by default settings). Proportions of CD45-positive cells were calculated as a cell weighted average (based on the total number of cells) for tissues with multiple non-contiguous tumor-enriched regions.

### Other pathologic features

Other pathologic features that were assessed in this cohort using available tissue include cellularity, stromal TILs (tumor infiltrating lymphocytes), HER2 FISH ratio, ER status, Her2 IHC, and Ki67 IHC, all assessed by board-certified pathologists. The Ki67 scoring, ER status, cellularity, TILs scoring, and cellularity measurement for this cohort was described previously^21^.

## Data Availability

Data Availability
Data associated with the manuscript to be available online.

## Data Availability

Data associated with the manuscript is available online: https://github.com/cancersysbio/BreastCancerSpatialProteomics

## Code Availability

Code associated with the manuscript is available online: https://github.com/cancersysbio/BreastCancerSpatialProteomics

## Acknowledgments

We thank NanoString for technical support. We thank members of the Curtis lab, especially Zheng Hu and Jose A. Seoane, for feedback on the manuscript and Joan Brugge for input on the HER2, and Ki67 IHC. This project was supported by awards to CC from the National Institutes of Health/National Cancer Institute (R01CA182514) and the Breast Cancer Research Foundation. C.C. is a Susan G. Komen Scholar. KLM is supported by F30CA239313-02 from the NIH/NCI. JLC was a Damon Runyon Physician-Scientist supported by the Damon Runyon Cancer Research Foundation (PST 11-17) and a Susan G. Komen Postdoctoral Fellowship Award (PDR17481769). MFP was supported by the Breast Cancer Research Foundation and Tower Cancer Research Foundation and R01CA182514. DJS and SS were also supported in part by R01CA182514.

## Author Contributions

KLM analyzed the data. RJ and EK contributed to statistical analyses. KLM, ZM, MK, ZZ, MH, JB contributed to data acquisition. GRB performed pathology review. JZ and MFP performed Ki67 and HER2 IHC. MFP performed HER2 FISH. SAH and DJS led the clinical trial and oversaw sample collection. KLM, JLC and CC interpreted the data. KLM and CC wrote the manuscript. CC conceived of and supervised the study. All authors read and approved the final manuscript.

---

**Extended Data 1.**
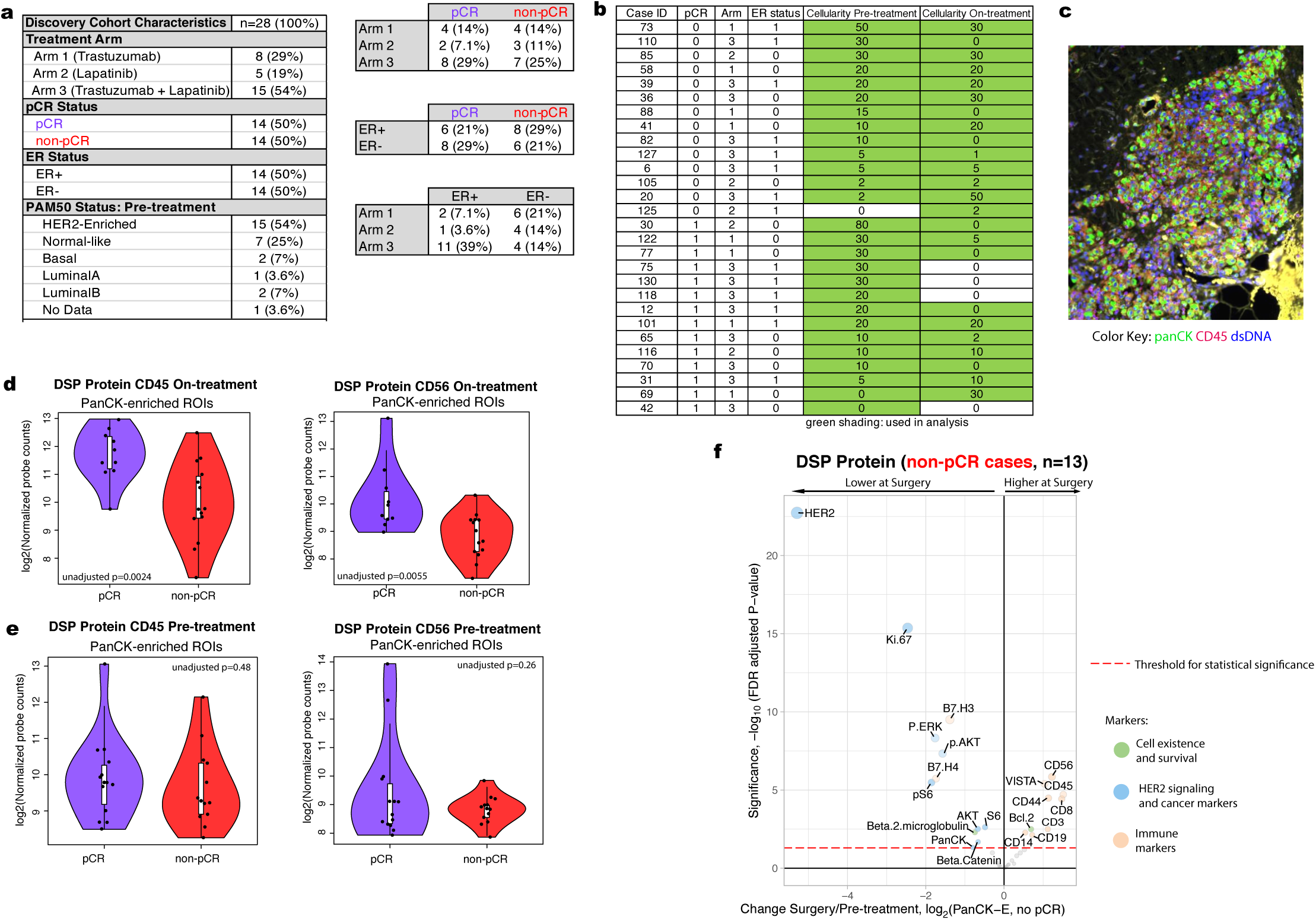
Discovery cohort description and preliminary proteomic analysis. a. Summary of the clinical characteristics of the TRIO-US B07 DSP discovery cohort, including treatment arm, pathologic complete response (pCR), estrogen receptor (ER) status, and PAM50 status inferred based on pre-treatment bulk expression data. Two-way contingency tables compare the distribution of ER status, pCR status, and treatment arm. b. Pathology-estimated cellularity pre-treatment and on-treatment for the discovery cohort. Samples with green shading indicate those used for subsequent analysis. For the pCR column, 0=non-pCR, 1=pCR. For ER status column, 0=ER-negative, 1=ER-positive. c. An example region from case 30 sampled on-treatment. While cellularity was estimated to be 0 based on pathology review of a distinct tissue section, tumor regions were identified upon imaging the tissue section used in this analysis. d. CD45 values and CD56 values from the Digital Spatial Profiling (DSP) protein data on-treatment in the pCR cases versus the non-pCR cases. Each point represents the average probe values for all panCK-enriched ROIs for that case On-treatment. The p-value was derived using a linear mixed-effect model over the multi-region data with blocking by patient. e. CD45 values and CD56 values from the protein DSP data pre-treatment in the pCR cases versus the non-pCR cases. For each violin plot, the white box represents the interquartile range and the black lines extending from the white box represent 1.5X the interquartile range. Analyses based on the discovery cohort. f. Volcano plot demonstrating treatment-associated changes from pre-treatment to surgery in tumors that did not undergo pCR using DSP protein expression levels in pancytokeratin-enriched (PanCK-E) regions. Significance, -log10(FDR adjusted p-value), is indicated along the y-axis.

**Extended Data 2.**
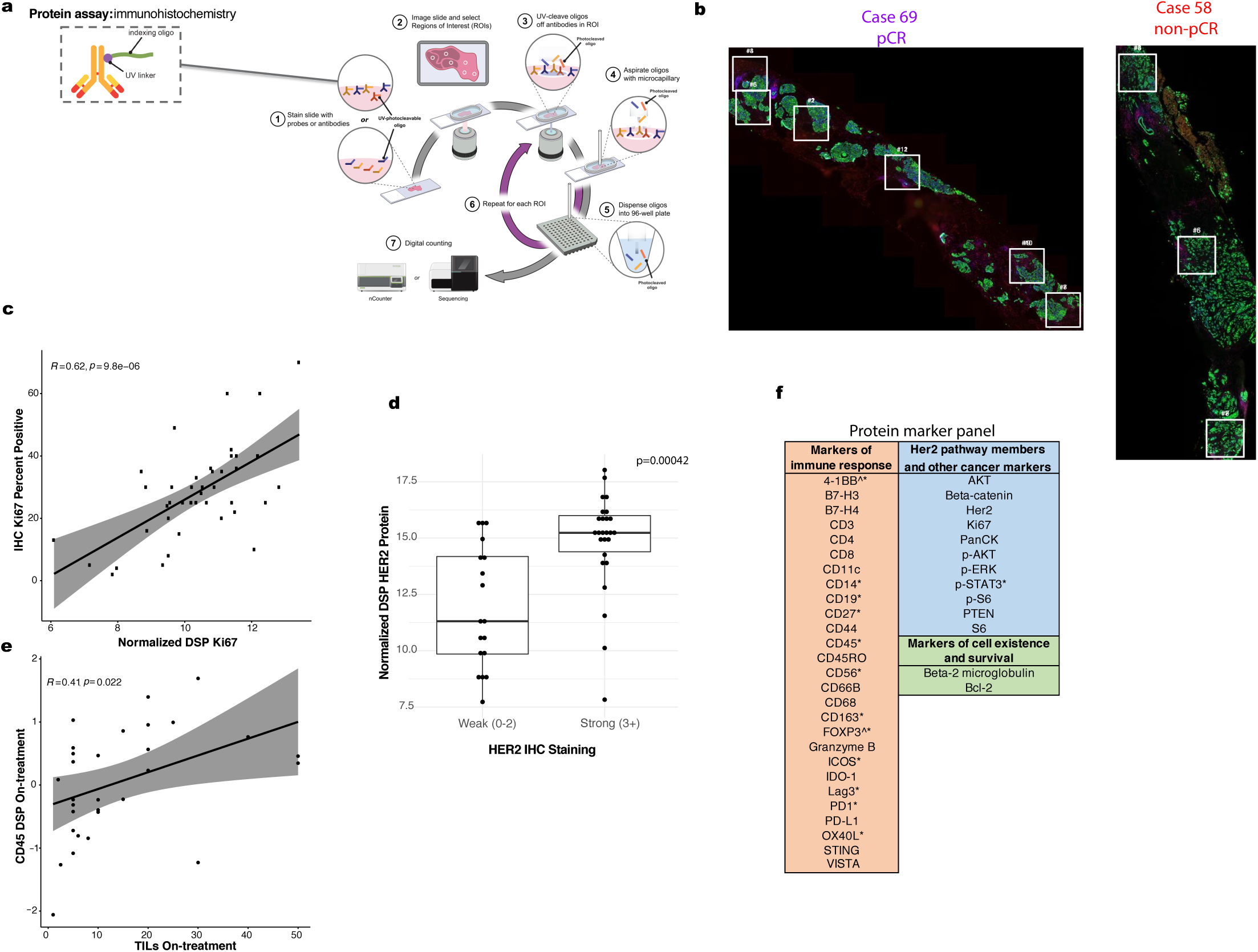
Digital Spatial Profiling (DSP) is used for multiplex protein quantification within tumor regions. a. NanoString DSP workflow summary. The slide is stained with the mix of protein antibodies. The antibodies have an indexing oligo attached, which is used for subsequent readout. ROIs (regions of interest) are selected and illuminated using UV (ultraviolet) light. The UV light causes the indexing oligos within the ROI to be cleaved off for collection and per-probe quantification. b. The location of spatially separated ROIs within tissue specimens for a representative pCR case (69) and a demonstrative non-pCR case (58). An average of 4 ROIs were profiled per tissue (range: 1-7). c. Correlation plot comparing Ki67 percent positive (evaluated using IHC) with normalized DSP Ki67 expression (averaged across all ROIs within a distinct tissue slice from the same case and timepoint). A total of 42 biopsies (24 pre-treatment and 18 on-treatment) with paired Ki67 IHC and DSP data were utilized in this analysis. Pearson correlation coefficient and corresponding p-value are also noted. d. Boxplot comparing normalized DSP Her2 expression (averaged across all ROIs from the same case and timepoint) between cases that exhibited strong (3+) IHC Her2 staining (using a distinct tissue slice from the same case and timepoint) or weaker (0-2) IHC Her2 staining. A total of 44 biopsies (23 pre-treatment and 21 on-treatment) with paired Her2 IHC and DSP data were utilized in this analysis. A Wilcoxon test was used to assess significance. e. Correlation plot comparing normalized on-treatment CD45 DSP protein values with on-treatment stromal tumor infiltrating lymphocyte (TILs) score for all cases with both data types available (n=31). f. Markers with a signal to noise ratio < 3 (Methods) in the discovery cohort indicated by a caret (^) and those with an SNR < 3 in the validation cohort are indicated with an asterisk (*).

**Extended Data 3.**
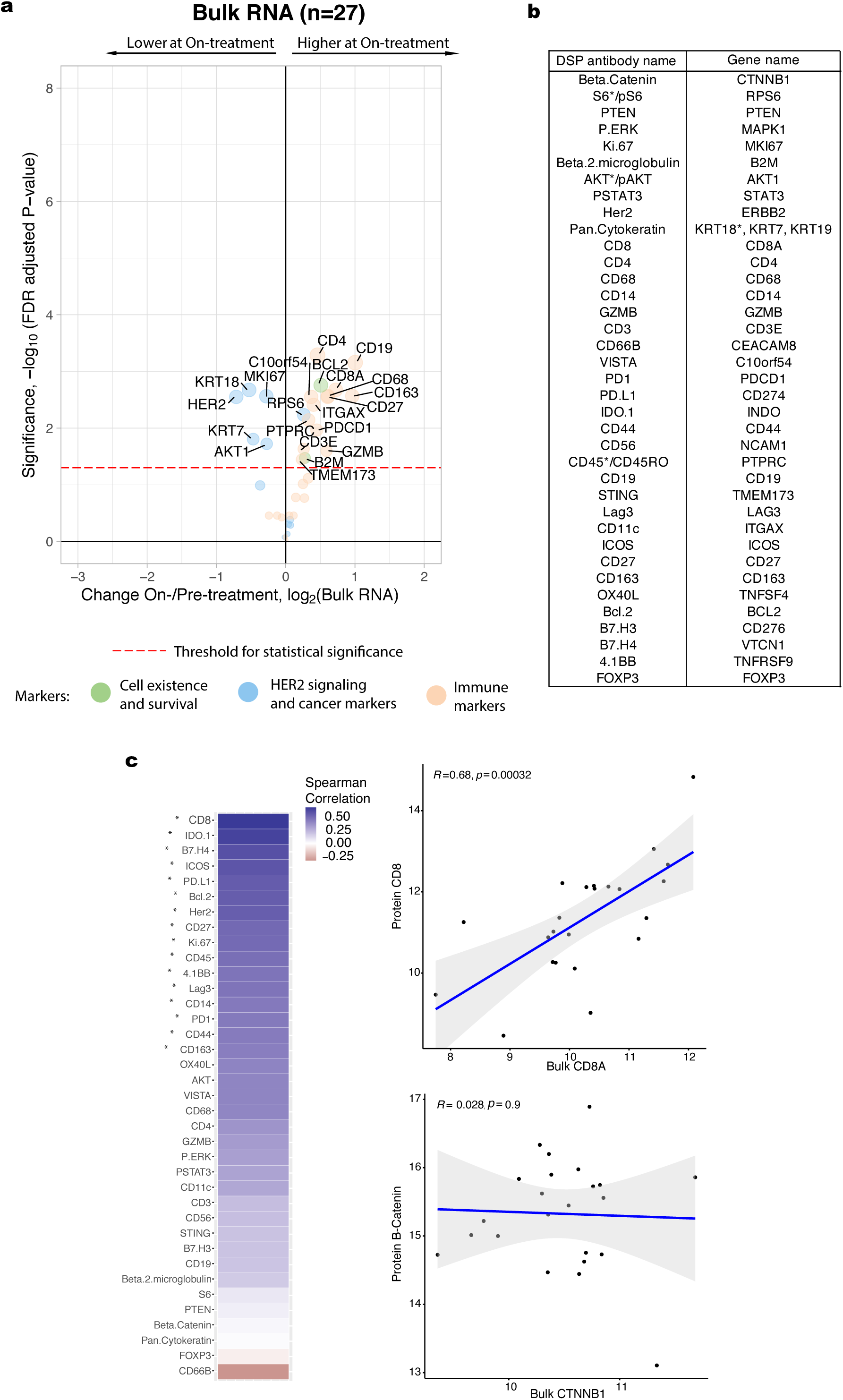
Treatment-associated changes observed with bulk RNA data. a. Volcano plot demonstrating treatment-associated changes based on comparison of pre-treatment versus on-treatment bulk RNA expression levels. RNA transcripts with corresponding Digital Spatial Profiling (DSP) protein markers were used in this analysis. Significance, -log10(FDR adjusted p-value), is indicated along the y-axis. Analyses based on the discovery cohort. b. Pairing of protein antibodies and gene names used in comparative analyses between DSP and bulk expression data. Genes listed here were used to generate the bulk expression volcano plot shown in panel a. For direct comparisons yielding the correlation plots shown in panel c, the marker names indicated by the asterisk were used; specifically, for proteins with multiple form (e.g. AKT/pAKT, CD45/CD45RO), the unmodified form of the protein was used when available and the breast-cancer associated keratin gene with the highest mean expression level was used. c. Spearman correlation between DSP protein probes (averaged across all ROIs per case) and bulk RNA transcripts corresponding to these markers pre-treatment. Significantly correlated probes (with p-value < .05) are indicated by an asterisk. Two exemplary correlation plots are shown, where each dot represents a single case. Analyses based on the discovery cohort.

**Extended Data 4.**
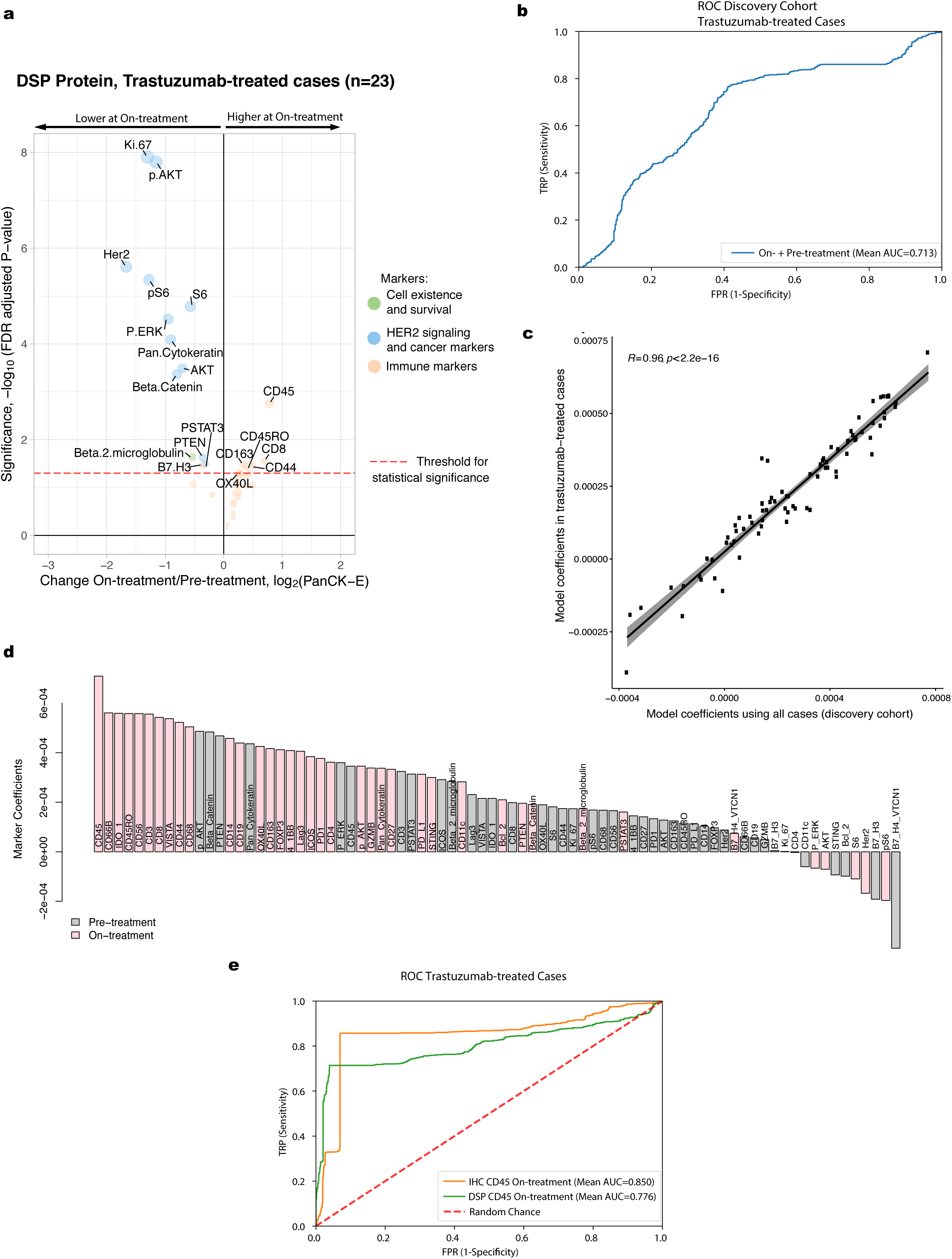
Treatment-associated changes and model performance assessed in cases treated with trastuzumab. a. Volcano plot demonstrating treatment-associated changes based on comparison of pre-treatment versus on-treatment protein marker expression levels in pancytokeratin-enriched (PanCK-E) regions in the trastuzumab-treated cases (arms 1 and 3, n=23). Significance, -log10(FDR adjusted p-value), is indicated along the y-axis. Analyses based on the discovery cohort. b. Receiver operating characteristic (ROC) curves for On-plus Pre-treatment DSP protein L2-regularized classifier in the discovery cohort using the subset of cases with data available at both timepoints that were treated with trastuzumab or trastuzumab+lapatinib (n=19). Model performance was assessed via cross-validation using the 40 DSP protein markers profiled in both cohorts c. Correlation plot comparing the marker coefficients for the On-plus Pre-treatment DSP protein trained using all cases in the discovery cohort and using only those cases treated with trastuzumab (arms 1 and 3). d. Coefficients for each marker in the L2-regularized On-plus Pre-treatment DSP protein model, trained using only those cases treated with trastuzumab (arms 1 and 3). e. OC curves for classifiers trained using either DSP on-treatment CD45 levels or IHC CD45 % positive. These models were evaluated on the subset of cases on which CD45 IHC was performed that were treated with trastuzumab or trastuzumab+lapatinib (n=20).

**Extended Data 5.**
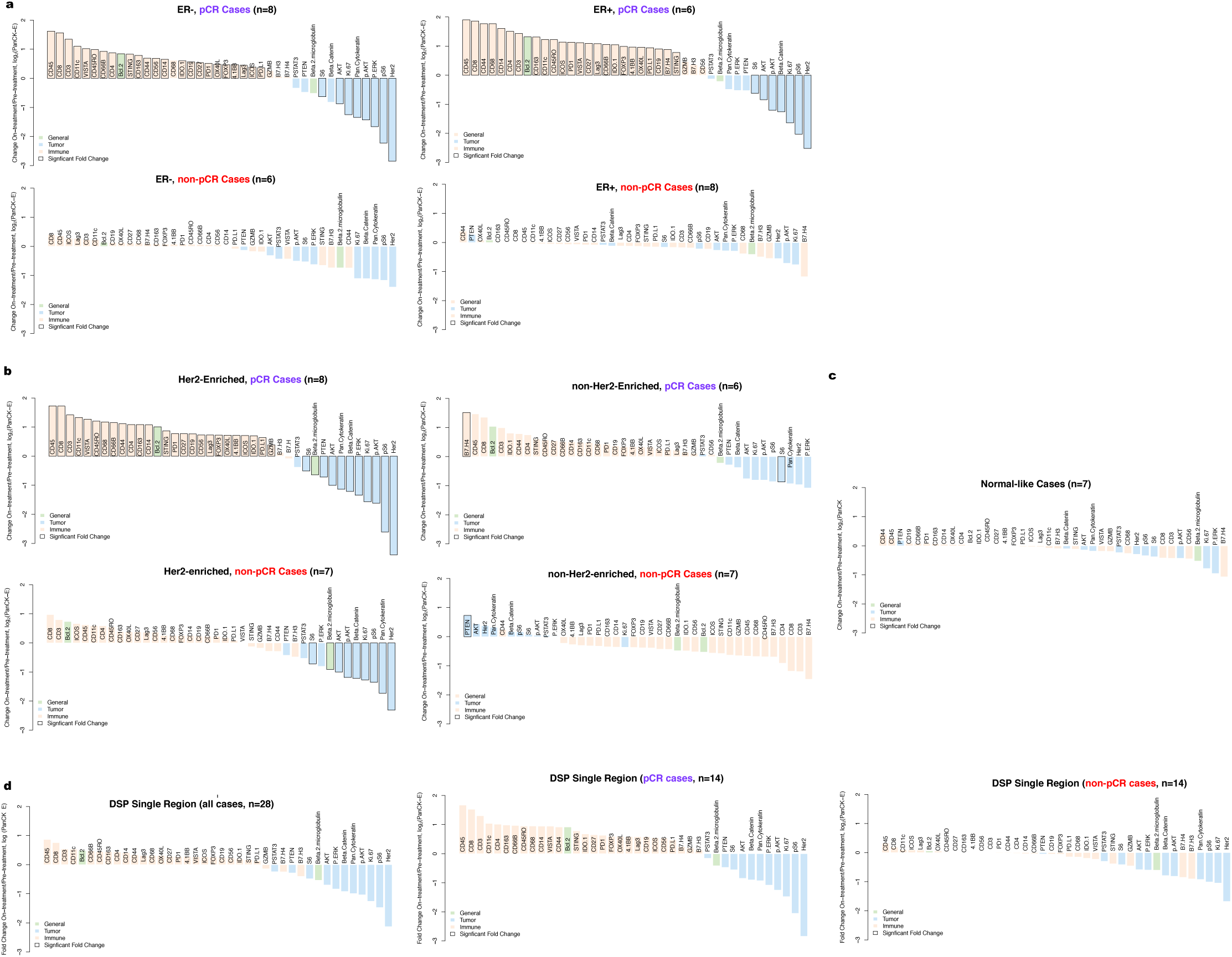
Waterfall plots illustrating treatment-associated changes in various data subsets. Waterfall plots show changes (pre-treatment to on-treatment) based on in pancytokeratin-enriched (PanCK-E) regions from DSP protein expression data. Analyses based on the discovery cohort. a. Input data was stratified both by estrogen receptor (ER) status and pathologic complete response (pCR) outcome. b. Samples were stratified both by PAM50 status (Her2-Enriched or other) and pCR. c. Waterfall plots illustrating treatment-associated changes (pre-treatment to on-treatment) in the Normal-like cases in the discovery cohort. PAM50 status was determined using bulk expression data from the pre-treatment biopsies. d. Waterfall plots when only one region is used to profile each sample (averaged across 100 iterations of random samples of a single region per timepoint), rather than the 2-7 regions from each sample used in other analyses. The leftmost plot is for all patients, and the plots on the right are stratified by pCR.

**Extended Data 6.**
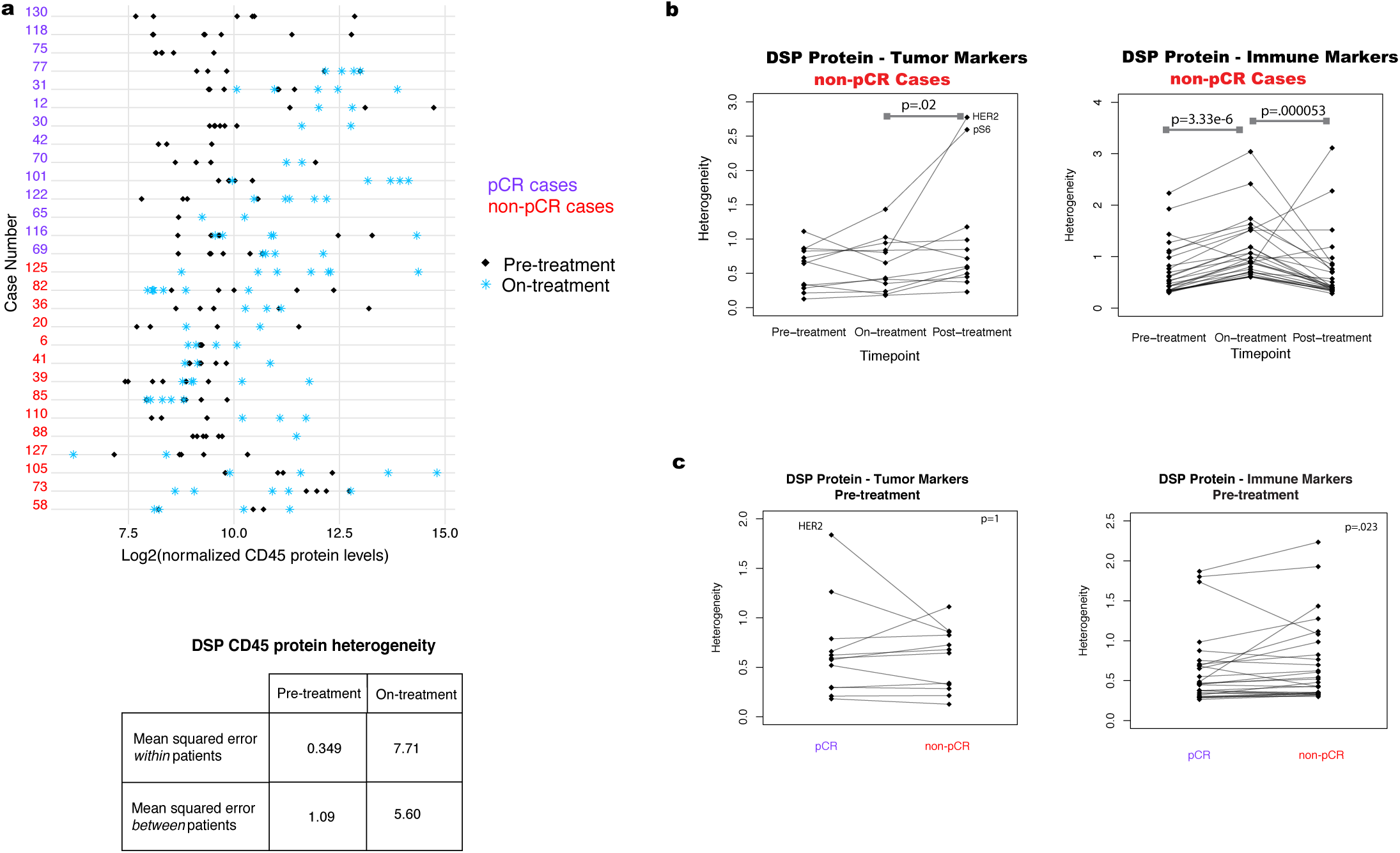
Regional heterogeneity profiled through treatment in tumor and immune markers. Analyses based on the discovery cohort. Heterogeneity was calculated as the mean squared error within patients based on analysis of variance. P-values are based on a two-sided Wilcoxon matched-pair signed rank test. a. Comparison of DSP CD45 protein levels pre-treatment and on-treatment for all regions profiled per case per timepoint. Also shown is a comparison of the mean squared error in DSP CD45 protein expression pre-treatment versus on-treatment within and between patients. b. Pre-, on-, and post-treatment heterogeneity for each DSP protein marker in non-pCR cases (patients with tumor cells present at surgery). c, Pre-treatment heterogeneity in DSP protein marker expression in pCR and non-pCR cases.

**Extended Data 7.**
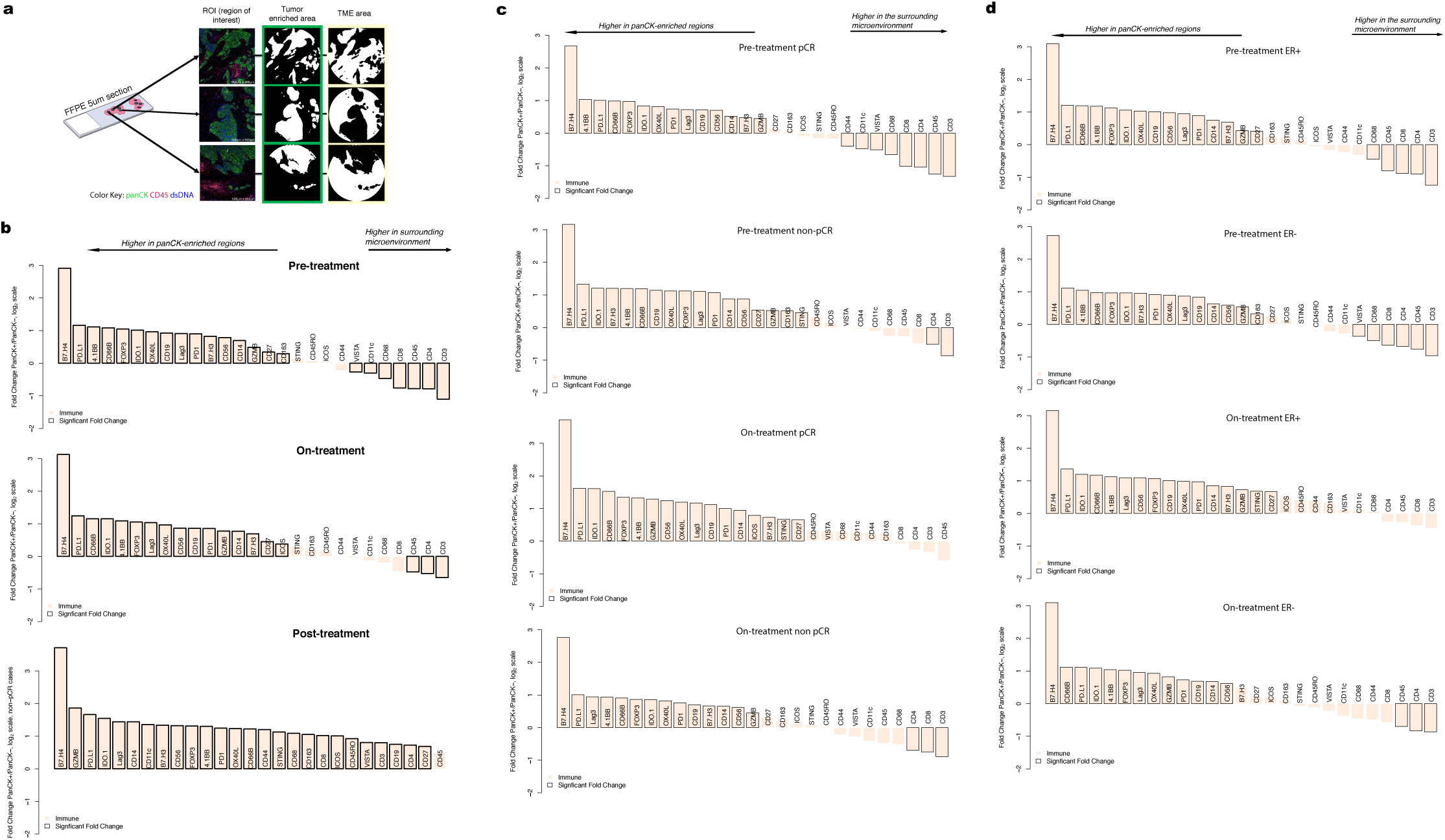
Comparison of immune markers in panCK-enriched and panCK-negative regions. a. Digital Spatial Profiling (DSP) was performed on multiple regions of interest (ROIs) per tissue sample. Protein counts were measured within phenotypic regions corresponding to the PanCK-enriched (tumor-enriched) masks that include tumor cells and co-localized immune cells and separately for the inverted mask corresponding to panCK-negative (tumor microenvironment, TME) regions. b-d. Waterfall plots, generated using the DSP protein data, comparing immune marker expression between the panCK-enriched regions and the surrounding panCK-negative regions. Analyses based on the discovery cohort. b. A comparison of all data pre-treatment on-treatment, and post-treatment. c. A comparison of pre-treatment and on-treatment timepoints, in pCR (n=14) and non-pCR cases (n=14). Pre-treatment, the correlation between immune marker fold-change values in the pCR and non-pCR cases was 0.98 indicating similar immune distribution across the panCK-enriched regions and surrounding microenvironment regardless of pCR outcome and this correlation remained high on-treatment (0.95). d. A comparison of pre-treatment and on-treatment timepoints, in ER-positive cases (n=14), and ER-negative cases (n=14).

**Extended Data 8.**
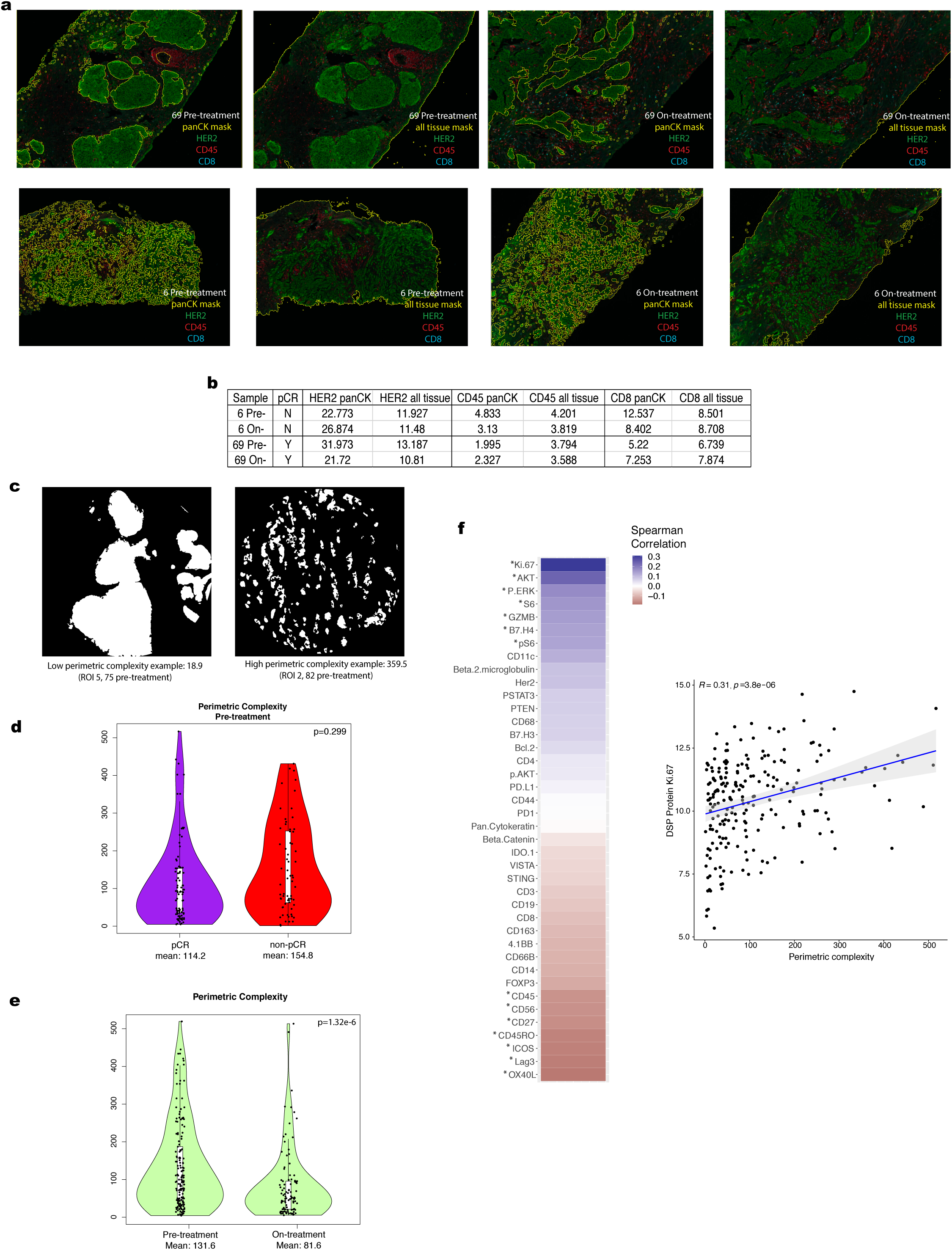
Multiplex immunohistochemistry and perimetric complexity. a. Multiplex immunohistochemistry (mIHC) images showing the distribution of HER2, CD45, and CD8 signal in representative tissue stamps pre-treatment and on-treatment. The panCK mIHC channel (not shown) was used to generate the panCK mask and the tissue mask (outlined in yellow). b. IHC marker expression levels for HER2, CD45, and CD8 were quantified for the whole tissue section (across all digitized sub-images) and within the panCK-enriched tumor regions (across all digitized sub-images). c. Illustration of panCK-enriched binary masks and perimetric complexity-based quantification of the tumor-microenvironment border. d. Comparison of perimetric complexity values pre-treatment between pCR cases and non-pCR cases. e. Comparison of pre-treatment versus on-treatment perimetric complexity values. PanCK-enriched ROIs were used to quantify perimetric complexity. P-values computed with a linear model, blocked by patient. f. Spearman correlation between the DSP protein expression values and perimetric complexity per region of interest (ROI) in the pre-treatment and on-treatment tissue specimens from the discovery cohort. Significantly correlated probes: p-value < .05 are denoted by an asterisk. Correlation plot for Ki-67, the marker with the highest correlation with perimetric complexity, where each dot represents an individual ROI. Analyses are based on the discovery cohort.

**Extended Data 9.**
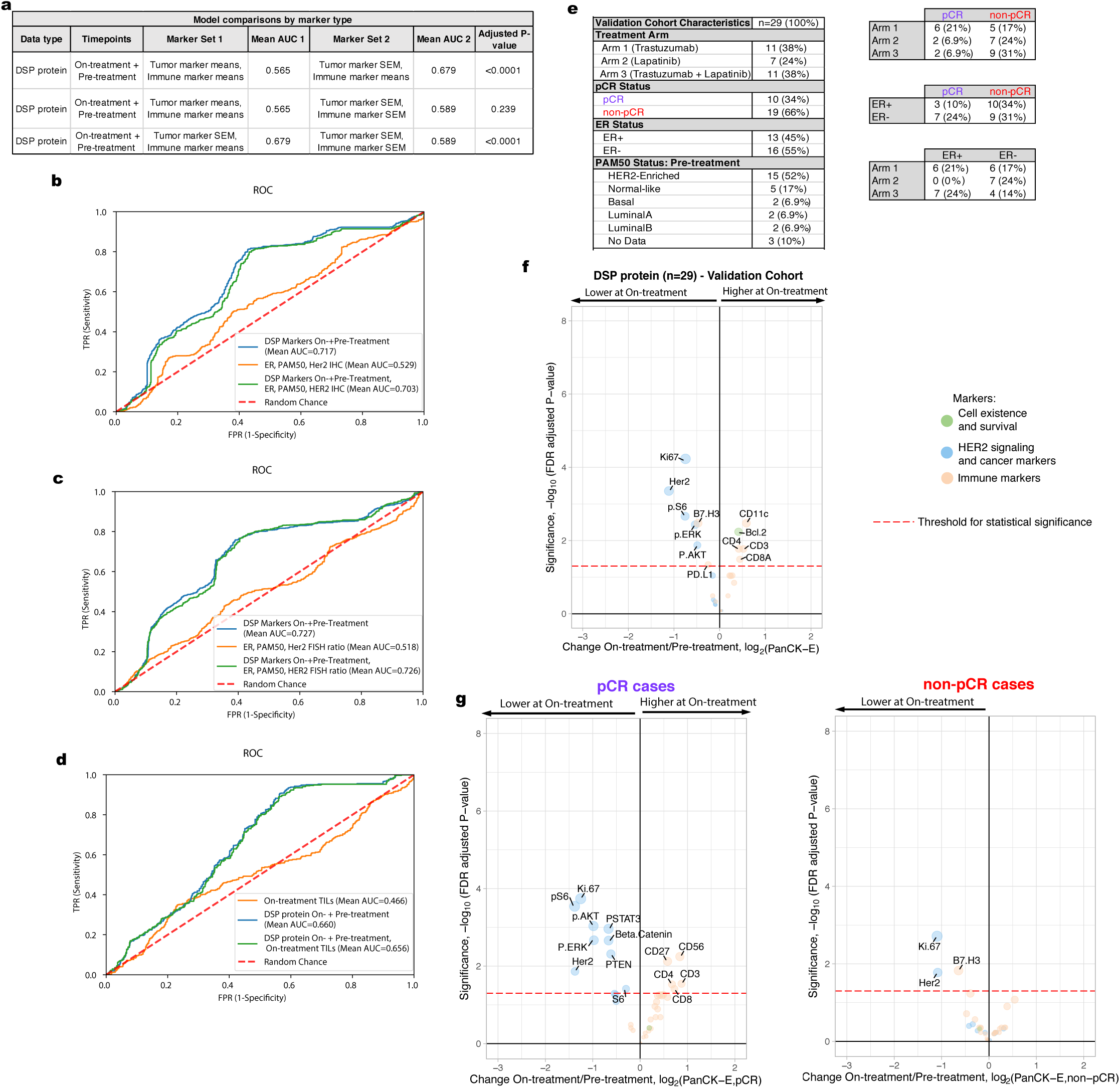
Comparison of alternative models including non-DSP measures and model validation cohort. a. AUROC (Area Under the Receiver Operating Characteristics) performance (using nested cross-validation with Holm-Bonferroni correction for multiple hypotheses) comparing DSP protein on-plus pre-treatment L2-regularized classifiers trained using marker means versus marker standard error of the mean (SEM) for tumor markers and immune markers. b-d. Receiver operating characteristic (ROC) curves and AUROC quantification for the On-plus Pre-treatment DSP protein L2-regularized classifier using all 40 markers compared to other models: (b) model trained using estrogen receptor (ER) status, PAM50 status, and HER2 IHC (3+ vs other) (n=19) and model incorporating these three measures as well as On-plus Pre-treatment DSP; (c) model trained using ER status, PAM50 status, and pre-treatment HER2 FISH ratio (n=21) and model incorporating these three measures as well as On-plus Pre-treatment DSP; (d) model trained using on-treatment stromal tumor infiltrating lymphocytes (TILs) (n=16) and model incorporating TILs as well as On-plus Pre-treatment DSP. Model comparisons were performed in the discovery cohort. e. Summary of the clinical characteristics for the TRIO-US B07 clinical trial Digital Spatial Profiling (DSP) validation cohort used for model testing. Treatment arm, pathologic complete response (pCR), ER status, and PAM50 status inferred based on pre-treatment bulk expression data are included. Two-way contingency tables compare the distribution of ER status, pCR status, and treatment arm. f. Volcano plot demonstrating treatment-associated changes based on comparison of pre-treatment versus on-treatment protein marker expression levels in pancytokeratin-enriched (PanCK-E) regions in the validation cohort. g. Volcano plots demonstrating treatment-associated changes in pCR versus non-pCR cases in the PanCK-E regions in the validation cohort. Significance, -log10(FDR adjusted p-value), is indicated along the y-axis.

**Extended Data 10.**
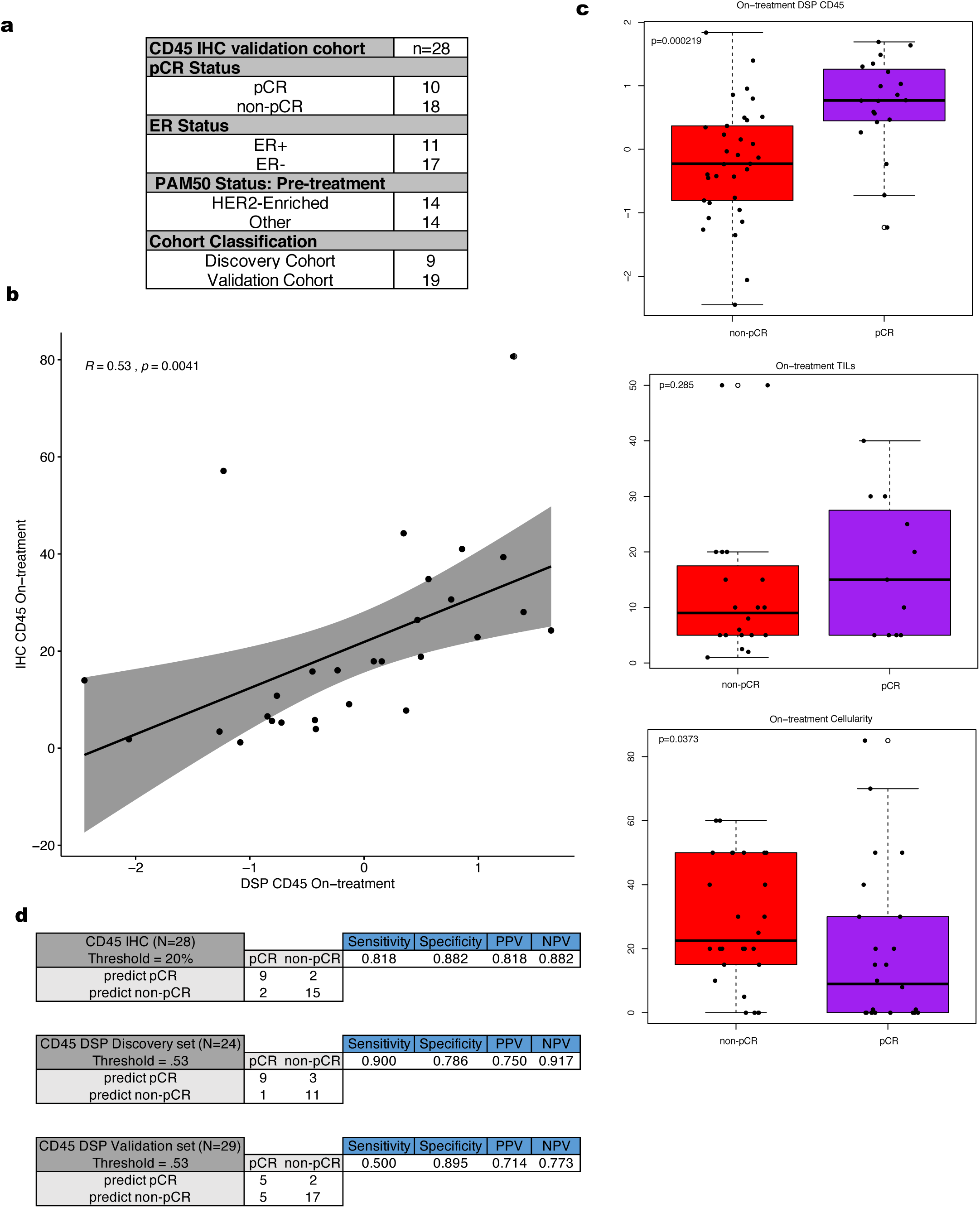
On-treatment immunohistochemistry for CD45 predicts pathologic complete response. a. Summary of the clinical characteristics of cohort used to evaluate IHC CD45 percent positive cells within tumor enriched regions as a biomarker. These features include pathologic complete response (pCR), estrogen receptor (ER) status, and PAM50 status. b. Correlation plot comparing the normalized on-treatment CD45 DSP protein values with the paired on-treatment CD45 % positive derived from IHC. c. Boxplots showing DSP on-treatment CD45 levels, on-treatment tumor cellularity, and on-treatment stromal tumor infiltrating lymphocyte (TILs) score stratified by pCR utilizing all available data where these features were measured across the discovery and validation cohorts. P-values were derived using the Wilcoxon rank-sum test. For each boxplot, the colored box represents the interquartile range and the black lines extending from the box represent 1.5X the interquartile range. d. For both the CD45 on-treatment IHC data and the normalized CD45 on-treatment DSP data, thresholds were defined to predict pCR vs non-pCR cases. Sensitivity, specificity, positive predictive value (PPV), and negative predictive value (NPV) are shown for such thresholds.

